# The genomic and transcriptional landscape of primary central nervous system lymphoma

**DOI:** 10.1101/2021.07.30.21261280

**Authors:** Josefine Radke, Naveed Ishaque, Randi Koll, Zuguang Gu, Elisa Schumann, Lina Sieverling, Sebastian Uhrig, Daniel Hübschmann, Umut H. Toprak, Cristina López, Xavier Pastor Hostench, Simone Borgoni, Dilafruz Juraeva, Fabienne Pritsch, Nagarajan Paramasivam, Gnana Prakash Balasubramanian, Matthias Schlesner, Shashwat Sahay, Debora Pehl, Helena Radbruch, Anja Osterloh, Agnieszka Korfel, Martin Misch, Julia Onken, Katharina Faust, Peter Vajkoczy, Dag Moskopp, Yawen Wang, Andreas Jödicke, Lorenz Trümper, Ioannis Anagnostopoulos, Dido Lenze, Michael Hummel, Clemens A. Schmitt, Otmar D. Wiestler, Stephan Wolf, Andreas Unterberg, Roland Eils, Christel Herold-Mende, Benedikt Brors, ICGC MMML-Seq Consortium, Reiner Siebert, Stefan Wiemann, Frank L. Heppner

**Author notes:** Corresponding authors: Josefine Radke, MD, Charité - Universitätsmedizin Berlin, Department of Neuropathologie, Charitéplatz 1 (Virchowweg 15), 10117 Berlin, Naveed Ishaque, PhD. ICGC MMML-Seq Consortium (full list of members of the ICGC MMML-Seq Consortium is listed in the Supplementary text). shared co-first authorship (Josefine Radke, Naveed Ishaque). shared senior authorship (Reiner Siebert, Stefan Wiemann, Frank L. Heppner).

## Abstract

Primary lymphomas of the central nervous system (PCNSL) are mainly diffuse large B-cell lymphomas (DLBCLs) confined to the central nervous system (CNS). Despite extensive research, the molecular alterations leading to PCNSL have not been fully elucidated. In order to provide a comprehensive description of the genomic and transcriptional landscape of PCNSL, we here performed whole-genome and transcriptome sequencing and integrative analysis of 51 lymphomas presenting in the CNS, including 42 EBV-negative PCNSL, 6 secondary CNS lymphomas (SCNSL) and 3 EBV+ CNSL and matched controls. The results were compared to an independent validation cohort of 31 FFPE CNSL specimens (PCNSL, n = 19; SCNSL, n = 9; EBV+ CNSL, n = 3) as well as 39 FL and 36 systemic DLBCL cases outside the CNS. Somatic genomic alterations in PCNSL mainly affect the JAK-STAT, NFkB, and B-cell receptor signaling pathways, with hallmark recurrent mutations including *MYD88* L265P (67%) and *CD79B* (63%), *CDKN2A* deletions (83%) and also non-coding RNA genes such as *MALAT1* (70%), *NEAT* (60%), and *MIR142* (80%). Kataegis events, which affected 15 of 50 identified driver genes and 21 of the top 50 mutated ncRNAs, played a decisive role in shaping the mutational repertoire of PCNSL. Compared to systemic DLBCL, PCNSLs exhibited significantly more focal deletions in 6p21 targeting the HLA-D locus that encodes for MHC class II molecules as a potential mechanism of immune evasion. Mutational signatures correlating with DNA replication and mitosis (SBS1, ID1 and ID2) were significantly enriched in PCNSL (SBS1: *p* = 0.0027, ID1/ID2: *p* < 1×10^−4^). Furthermore, *TERT* gene expression was significantly higher in PCNSL compared to ABC-DLBCL (*p* = 0.027). Although PCNSL share many genetic alterations with systemic ABC-DLBCL in the same signaling pathways, transcriptome analysis clearly distinguished both into distinct molecular subtypes. EBV+ CNSL cases may be distinguished by lack of recurrent mutational hotspots apart from IG and *HLA-DRB* loci.

## Introduction

Central nervous system (CNS) lymphomas are predominantly aggressive neoplasms involving brain, meninges, spinal cord and eyes^1,2^. Two clinical subtypes of CNSL can be distinguished: primary central nervous system lymphoma (PCNSL), which is confined to the CNS; and secondary central nervous system lymphoma (SCNSL) presenting initially with systemic, non-CNS involvement. SCNSL reflects a spread of a peripheral lymphoma to the CNS and its presentation, tropism, outcome and therapeutic options differ from PCNSL^3,4^.

PCNSL incidence is increased in immunocompromised patients, in which the tumor cells are typically Epstein-Barr virus (EBV)-positive. In contrast, PCNSL in immunocompetent patients is typically EBV-negative. The mechanisms leading to the topographical restriction of PCNSL are still matter of scientific debate^5^. PCNSL is classified as diffuse large B-cell lymphoma (DLBCL) in the vast majority of cases (approx. 90%) which immunohistochemically most often show a non-GCB immunophenotype^1,6,7^ according to the Hans classification^8^. The tumor cells express pan B-cell markers (CD19, CD20, and CD79a), the germinal center (GC)-associated molecule BCL6^9^, and the post-GC-associated marker MUM1/IRF4^10^. By gene expression profiling, the tumor cells are most closely related to late germinal center (exit) B-cells^11^. Pathomechanistic genomic alterations involving Toll-like- and B-cell receptor (TLR, BCR) signaling pathways have been identified in previous studies revealing a very high frequency of somatic nonsynonymous mutations in genes such as *MYD88, CARD11*, and *CD79B*^12-16^. Additionally, often homozygous HLA class II^17,18^ and *CDKN2A* loss, recurrent *BCL6* translocations^19,20^ and structural variants at chromosome band 9p24.1 (affecting *CD274*/*PD-L1 and PDCD1LG2/PD-L2*)^21^ as well as *TBL1XR1* variants^22^ have been repeatedly described in PCNSL^23,24^. These mutational patterns suggest PCNSL to be genetically similar to recently described “MCD”, “C5” or “MYD88-like” subtypes for which a derivation from long-lived memory B-cells has been proposed^25-30^.

The outcome of PCNSL, even in immunocompetent hosts, is poor compared to most primary systemic DLBCL^31^, though probably not worse than that of DLBCL of the MCD/C5 group in general^26^. High-dose methotrexate (MTX) remains the commonly administered therapy but the use of rituximab (monoclonal anti-CD20 antibody) has been shown to be effective^32,33^. However, reports on rituximab efficiency in PCNSL are conflicting^34-36^. Genomic studies have suggested lymphoma cell proliferation and survival to be driven at least in part, by deregulated TLR, BCR, JAK-STAT, and NFkB signaling pathways inducing constitutive NFkB activation^37-39^. Therefore, inhibitors up- and downstream of NFkB such as ibrutinib, known to inhibit Bruton’s tyrosine kinase (BTK) as critical mediator of B-cell receptor signaling, and lenalidomide which was shown to have indirect effects on tumor immunity have been applied, which seem to be effective therapeutic alternatives in PCNSL^40-43^. PD-L1/2 blockade is discussed as another therapeutic option^44^.

Despite all progress in the molecular characterization of PCNSL in the last decades, our understanding of the genetic and transcriptional alterations of PCNSL is by far not comprehensive. The few previous next generation sequencing (NGS) studies of PCNSL have been limited to target enrichment only of exons, or whole-genome analysis of very few samples^38,45-50^. Therefore, pathogenic mechanisms other than coding variants, such as non-coding and regulatory changes, structural variants or mutational mechanisms related to the genome-wide distribution of somatic hypermutation (SHM) have not been fully elucidated in PCNSL. Unbiased omics profiling, such as whole-genome sequencing (WGS) studies integrated with transcriptome sequencing, are currently the methods of choice to illuminate the role of non-coding mutations^51,52^. In addition, these approaches can unravel various molecular mechanisms deregulating driver genes in PCNSL, which are necessary for diagnosis, risk stratification, and treatment in the era of precision and targeted therapies. Thus, we here performed whole-genome and transcriptome sequencing in 51 B-cell lymphomas presenting in the CNS, including 42 PNCSL samples from immunocompetent patients, to comprehensively describe the mutational and transcriptonal landscape of PCNSL.

## Material and Methods

### CNS lymphoma (CNSL) study cohort

Fresh frozen and paraffin embedded primary central nervous system lymphoma (PCNSL) and secondary central nervous system lymphoma (SCNSL) tumor tissue and matching blood samples (germline control) were acquired from the Department of Neuropathology, Charité, Berlin (Germany), and the Department of Neurosurgery, Heidelberg (Germany) from chemotherapy-naïve patients. All procedures performed in this study were in accordance with the ethical standards of the respective institutional research committees and with the 1964 Helsinki declaration and its later amendments or comparable ethical standards. The Charité ethics committee (Charitéplatz 1, 10117 Berlin, Germany) approved the study (EA1/245/13). Age at diagnosis, tumor localization, peripheral manifestation (bone marrow biopsy result, CT/MRI scan), first line therapy, follow-up as well as overall survival (OS) in months were evaluated. Additional control samples from age-matched, postmortem, and non-neoplastic brain (n = 2) were analyzed. The diagnosis was confirmed by at least two experienced (neuro)pathologists. The morphologic characteristics were evaluated on both, the fresh-frozen (FF) and formalin-fixed and paraffin-embedded (FFPE) tissue sections of the tumor biopsies. The tumor cell content in the cryopreserved sample material was estimated to be at least 60% based on pathological evaluation. Basic immunophenotypic characterization was performed on FFPE tissue sections (**Supplementary figure 1 A**) of the diagnostic tumor biopsies using an immunohistochemical panel including antibodies directed against CD20, CD10, BCL6, CD3, Ki67, MUM1/IRF4 and EBV (LMP1). To further exclude an EBV association, all cases with indefinite EBV IHC were investigated applying Epstein Barr virus (EBV) PCR as previously described^53^ (**Supplementary figure 1 B**). For the categorization of GCB and non-GCB, the samples were stratified according to the Hans classification^8^ (CD10, BCL6, MUM1. We enrolled CNSL from a total of 51 patients for whole-genome (WGS, n = 38) and RNA sequencing (RNAseq, n = 37), including n = 24 samples subjected to both workflows. The study cohort and sample size as well as the experimental design, analysis workflow, diagnosis, and quality metrics of WGS and RNAseq are displayed in **Figure 1** and **Supplementary table 1**. The inclusion criteria were based on the diagnosis of PCNSL and SCNSL according to the recent WHO classifications of tumors of hematopoietic and lymphoid organs and tumors of the central nervous system^1,2,7,54,55^.

**Figure 1:**
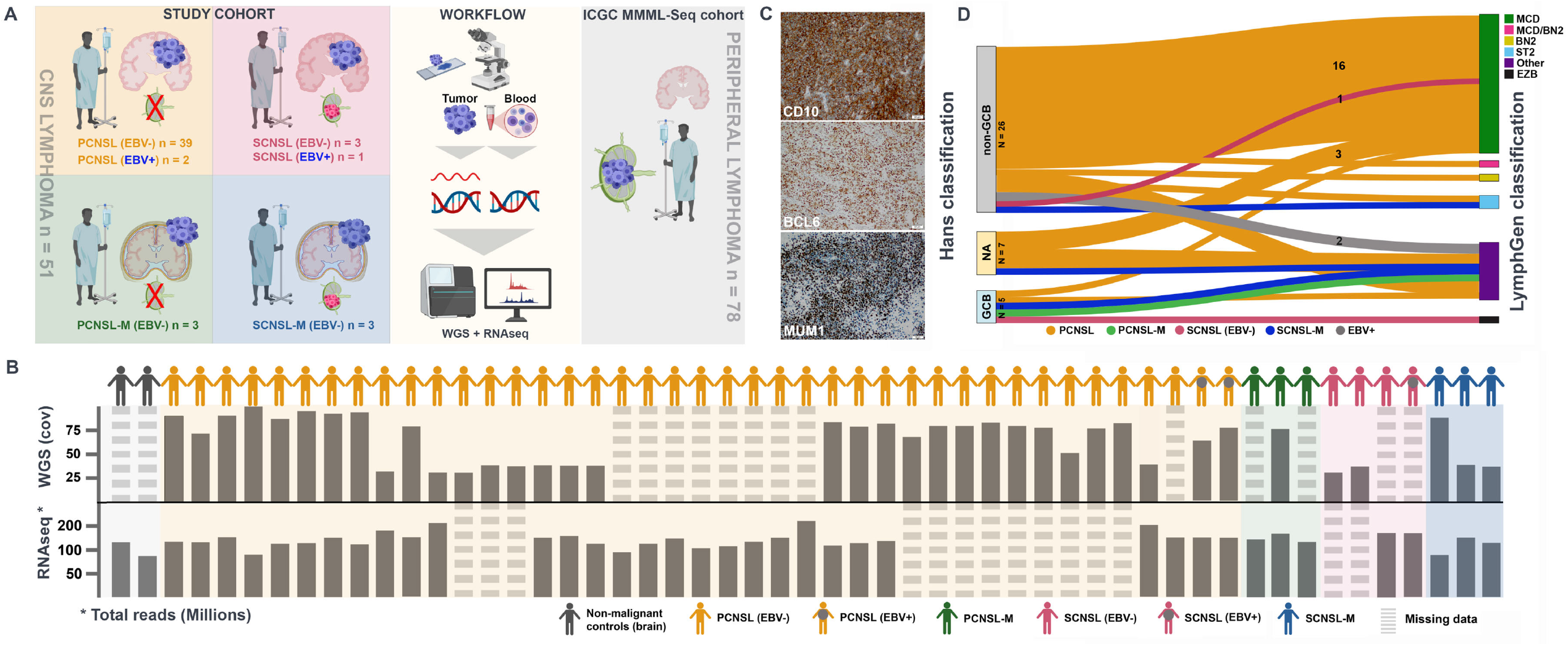
Study design and multi-omic analysis of the CNSL cohort. Panel A demonstrates the study design, study cohort along with sample size, and sequencing approach (**A**). The central nervous system lymphoma (CNSL) cohort consists of 51 primary and secondary lymphoma (PCNSL and SCNSL) patients. Whole-genome sequencing was performed using tumour tissue and matched peripheral blood samples. For RNA sequencing, we additionally analysed normal controls (non-malignant brain tissue (frontal lobe)) and included data from peripheral lymphomas without CNS manifestation for validation (ICGC MMML-seq cohort). The lower panel (**B**) lists all CNS lymphoma derived from PCNSL or SCNSL patients as well as controls. The depth of coverage (cov) for whole-genome sequencing and total number of reads in RNA sequencing are given for each tumour sample. Whole-genome sequencing data was obtained from n=38 PCNSL/SCNSL patients. RNA sequencing data was generated from n=37 PCNSL/SCNSL patients and n=3 normal controls. In 24 PCNSL/SCNSL cases, we obtained patient-matched whole-genome and RNA sequencing data. Striped bars indicate data sets missing for individual samples. The CNSL specimen were examined according to the Hans classification (CD10, BCL6, and MUM1 (**C**)). Additionally, we classified all n=38 whole-genome sequencing CNSL samples according to the genetic DLBCL subtypes using the LymphGen algorithm as described by Wright *et al*., 2020 (**D**). The results are displayed in a Sankey plot.

### ICGC MMML-Seq Consortium samples

For comparison, we used and reanalyzed an early release of meanwhile published whole-genome and RNA sequencing data obtained by the ICGC MMML-Seq Consortium from systemic diffuse large B-cell lymphoma (DLBCL, total: n = 36, WGS: n = 29, RNAseq: n = 36, both workflows: n = 29), follicular lymphoma (FL, total: n = 39, WGS: n = 39, RNAseq: n = 38, both workflows: n = 38), and one “double hit” (DH)-lymphoma with a molecular BL signature^30^. In addition, we included WGS and RNAseq data from a single EBV-PCNSL case as well as RNAseq data from two nodal marginal zone lymphomas (nMZL) as well as naïve (n = 5) and GC B-cells (n = 5) as normal controls^56^. These data were obtained by the ICGC MMML-Seq consortium in accordance to protocols previously published^51,52^.

### DNA and RNA isolation

DNA and RNA were obtained from fresh frozen CNSL tumor samples. RNA and genomic DNA were isolated from 15 to 30 10 μm cryosection slices (depending on the tissue size). DNA from tumor samples and their matched blood controls was isolated according to standard procedures. Total RNA from tumor samples was extracted using the RNeasy® Plus Mini Kit (Qiagen, Hilden, Germany) according to the manufacturer’s instructions. The RNA integrity number (RIN) was determined using an Agilent 2200 TapeStation system (Agilent Technologies, Santa Clara, CA).

### Whole-genome sequencing and data processing

#### Whole-genome sequencing

The DNA libraries of the tumor and matched control samples were prepared according to the Illumina TruSeq Nano DNA Library protocol using the TruSeq Nano DNA library Preparation Kit (Illumina, Hayward, CA; estimated insert size of 350 bp). Paired-end sequencing was performed on Illumina HiSeq X (2 x 150 bp) instruments using the TruSeq SBS kit, Version 3.

#### Alignment of sequencing reads

Sequencing reads were aligned using the DKFZ alignment workflow from ICGC Pan-Cancer Analysis of Whole Genome projects (https://dockstore.org/containers/quay.io/pancancer/pcawg-bwa-mem-workflow). Briefly, read pairs were mapped to the human reference genome (build 37, version hs37d5) using bwa mem (version 0.7.8) with minimum base quality threshold set to zero [-T 0] and remaining settings left at default values^57^, followed by coordinate sorting with biobambam bamsort (version 0.0.148) with compression option set to fast (1) and marking duplicate read pairs with biobambam bammarkduplicates with compression option set to best (9)^58^.

#### Small mutation calling and annotation

Somatic small variants (SNVs and indels) in matched tumor normal pairs were called using the DKFZ in-house pipelines as previously described^59^. Briefly, the SNVs were identified using samtools and bcftools version 0.1.1957^60^ and then classified as somatic or germline by comparing the tumor sample to the control, and later assigned a confidence which is initially set to 10, and subsequently reduced based on overlaps with repeats, DAC blacklisted regions, DUKE excluded regions, self-chain regions, segmental duplication records as introduced by the ENCODE project^61^ and additionally if the SNV exhibited PCR or sequencing strand bias. Only SNVs with confidence 8 or above were considered for further analysis. Tumor and matched blood samples were analyzed by Platypus^62^ to identify indels. Indel calls were filtered based on Platypus internal confidence calls, and only indels with confidence 8 or greater were used for subsequent analysis. In order to remove recurrent artifacts and misclassified germline events, somatic indels that were identified as germline in at least two patients in the CNS lymphoma cohort were excluded.

The protein coding effect of somatic SNVs and indels from all samples were annotated using ANNOVAR^63^ according to GENCODE gene annotation (version 19) and overlapped with variants from dbSNP10 (build 141) and the 1000 Genomes Project database. Mutations of interest were defined as somatic SNV and indels that were predicted to cause protein coding changes (non-synonymous SNVs, gain or loss of stop codons, splice site mutations, and both frameshift and non-frameshift indels), and also synonymous exonic mutations on non-coding genes.

#### Tumor in normal contamination detection

We applied the TiNDA (tumor in normal detection algorithm) workflow to account for potential tumor in normal contamination leading to false negative calls as previously described^59^. Briefly, the B-allele frequency (BAF) was calculated from the tumor and control samples. Positions overlapping with common variants were filtered out. Then, the clustering algorithm from Canopy^64^ was applied to the BAF values for the positions in tumor vs control using a single pass run, assuming 9 clusters. The clusters that were determined to be tumor-in-normal had to have 75% of positions above the identity line (where the VAF in the tumor sample is the same as the VAF in the control sample). These identified mutations were then reclassified as somatic instead of the original germline annotation. All but 4 CNSL WGS samples exhibited evidence for tumor in normal. On average 31 SNVs (range 0-136) were “rescued” in PCNSL, 6 in PCNSL-M (6-6, single sample), 27 (19-34) in SCNSL, 22 (9-43) in SCNSL-M, and 0 in the EBV-positive sub-cohorts (0-0, 2 samples). In total, only 6 SNVs with protein coding effects were rescued, including the MYD88 p.L265P mutation in sample LS-0102, which had 3 of 47 read support in the control, and 86 of 170 reads supporting the variant in the tumor sample (**Supplementary table 2**).

#### Genomic structural rearrangements

Genomic structural rearrangements (SVs) were detected using SOPHIA v.34.0^65^. Briefly, SOPHIA uses supplementary alignments as produced by bwa-mem as indicators of a possible underlying SV. SV candidates are filtered by comparing them to a background control set of sequencing data obtained using normal blood samples from a background population database of 3261 patients from published TCGA and ICGC studies and both published and unpublished DKFZ studies, sequenced using Illumina HiSeq 2000, 2500 (100 bp) and HiSeq X (151 bp) platforms and aligned uniformly using the same workflow as in this study. Gencode V19 was used for the gene annotations. We used the script draw_fusions.R from the Arriba package^66^ to visualize SVs generated by SOPHIA.

#### Copy number alterations and allelic imbalances

Allele-specific copy-number aberrations were detected using ACEseq (allele-specific copy-number estimation from whole-genome sequencing)^67^. ACEseq determines absolute allele-specific copy numbers as well as tumor ploidy and tumor cell content based on coverage ratios of tumor and control as well as the B-allele frequency (BAF) of heterozygous SNPs. SVs called by SOPHIA were incorporated to improve genome segmentation.

Final copy number segments were further smoothed to calculate the total number of gains and losses. Neighboring segments were merged if they rounded to the same copy number and deviated by less than 0.5 copies in case of segments <20 kb or deviated by less than 0.3 copies otherwise. Remaining segments <500 kb were merged with their closer neighbor based on allele-specific and total copy number and once again segments smaller than 2 Mb deviating by less than 0.4 copies were merged. Based on the resulting segments the number of gains and losses was estimated.

Furthermore, the fraction of aberrant genome was calculated as the fraction of the genome that is classified either as duplication or deletion (>0.7 deviation from the ploidy) or was identified as a loss of heterozygosity.

#### Classification of mutational hotspots (kataegis events)

Mutational hotspots indicating putative kataegis events (likely due to somatic hypermutation (SHM) or aberrant SHM) were defined as regions with at least 6 somatic SNVs within an average intermutational distance of 1000 bp or less, as previously used by Alexandrov and colleagues^68^. A gene was described to be targeted by kataegis if its definition (from Gencode version 19 gene models) overlapped with at least 1 kataegis region in at least 1 sample. While many of these kataegis loci are indeed SHM/aSHM targets, located 2.5 kb from the transcription start site (TSS), we cannot completely control for all PCNSL-specific TSSs due to the normal brain background tissue.

#### Mutational signatures

Supervised mutational signature analysis was performed using YAPSA development version 3.13^69^ using R 4.0.0. Briefly, the linear combination decomposition (LCD) of the mutational catalog with known and predefined PCAWG COSMIC signatures^70^ was computed by non-negative least squares (NNLS). The mutational signature analysis was applied to the mutational catalogs for SNVs (or single bas substitutions, SBS) and indels of all tumor samples. Signature-specific cutoffs were applied and cohort level analysis was used for detecting signatures as recommended by Huebschmann *et al*^*30*^. The cutoff used corresponds to “cost factors” of 10 for SNVs and 3 for indels in the modified ROC analysis.

#### Integration of different variant types

SNVs, indels, SVs and CNAs were integrated in order to account for all variant types in the recurrence analysis. All genes with SNVs or indels in coding regions (nonsynonymous, stop gain, stop loss, splicing, frameshift and non-frameshift events) and ncRNA (exonic) were included. Any SV with breakpoints directly lying on a gene (SV direct) were considered for oncoprints, however SVs were also annotated to a gene when they were either within 100kb of a gene (SV near), or to the closest gene (SV close) for SV recurrence analysis to account for regulatory mutations such as enhancer hijacking events. Genes were annotated with CNAs if they were completely or partially affected. Chromosome level CNVs events were determined once >30% of a chromosome arm was altered. Only focal CNA events were taken into account for variant integration, as these are more likely to target specific genes within the affected region than large events such as whole chromosome arm events. To capture the precise target of CNVs, we employed results from GISTIC. Finally, genes affected by SNVs, indels, directly hit by SVs or genes with focal CNAs were considered for the recurrence analysis and added to the oncoprints.

#### Mutual exclusivity and inclusivity analysis

Mutual exclusivity analysis was performed to investigate the relationship between *MYD88* mutations with other implicated drivers from the IntOGen analysis including SNVs, indels, SVs, CNAs. The minimal recurrence threshold was set to 5. We applied the commonly used Fisher’s exact test and the CoMET test^71^ for both co-occurrence and mutual exclusivity. Fisher’s right tailed test was used to support co-occurrence when the number of samples with alterations in both genes is significantly higher than expected by chance. Additionally, Fisher’s left tailed test was used to suggest mutual exclusivity when the number of samples with alterations in both genes is significantly lower than expected. Resultant *p*-values were corrected for multiple testing by FDR.

#### Mutational significance analysis

The IntOGen pipeline^72^ algorithm was applied to identify significant cancer drivers in the core set of PCNSL samples (n=30) based on the hg19 genome assembly. IntOGen v 3.0.4 was installed via conda from the bbglab anaconda channel. The relevant conda environment setup included explicit definitions of python v3.5.5 (with libraries pandas v0.17). In addition, a local installation of perl v5.16.3 was used, with installation of perl libraries Digest-MD5 v2.58 via cpan and perl-DBI v1.627-4.el7.x86_64 via yum package managers. The background intogen database (bgdata) was automatically downloaded using the command ‘intogen --setup’ which downloaded the 20150729 background databases. The IntOGen run specific parameters included running on 4 cores, Matlab Compiler Runtime v8.1 (2013a) and MutSigCV v1.4. Significance thresholds of 10% FDR were used for oncodrivefm, oncodriveclust and mutsig. Sample thresholds of 2 and 5 were used for oncodrivefm and oncodriveclust respectively. IntOGen reported 50 genes to be significant drivers.

#### Telomere content estimation

The telomere content was determined from whole-genome sequencing data using the software tool TelomereHunter using default settings (filtering of telomere reads: at least 6 telomere repeats per 100 bp read length)^73^. Briefly, unmapped reads or reads with a very low alignment confidence (mapping quality lower than 8) containing six non-consecutive instances of the four most common telomeric repeat types (TTAGGG, TCAGGG, TGAGGG, and TTGGGG) were extracted. The telomere content was determined by normalizing the telomere read count to all reads in the sample with a GC-content of 48– 52%. In the case of tumor samples, the telomere content was further corrected for the tumor purity (as estimated by ACEseq) using the following formula:

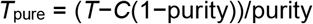

Here, *T* and *C* are the telomere contents of the tumor and control sample, and *T*_pure_ is the purity-corrected telomere content of the tumor sample.

#### LymphGen classification

All WGS samples were classified according to the LymphGen algorithm described by Wright *et al*.^27^ which categorises DLBCL samples into the different genetic subtypes MCD, N1, A53, BN2 ST2, EZB (MYC+ and MYC-), based on genetic aberrations in subtype predictor genes. The algorithm requires information on mutations, copy-number alterations, and fusions. According to Wright et al. ^27^, the results are displayed in **Supplementary table 1**. For a detailed description of the method, please refer to the **Supplementary information**.

### RNA sequencing and data processing

#### RNA library preparation and sequencing

RNA libraries of the tumor samples and normal brain samples were prepared using the TruSeq RNA library preparation Kit Set A and B, following the manufacturer’s instructions at an insert size of ∼300 bp. Two barcoded libraries were pooled per lane and sequenced on Illumina HiSeq2000 or HiSeq4000 platforms.

#### RNAseq alignment and expression quantification

RNAseq reads were aligned and gene expression quantified as previously described^74^. Briefly the RNAseq read pairs were aligned to the STAR index generated reference genome (build 37, version hs37d5) using STAR in 2 pass mode (version 2.5.2b)^74,75^. Duplicate reads were marked using sambamba (version 0.4.6) and BAM files were coordinate sorted using SAMtools (version 0.1.19). featureCounts (version 1.5.1)^76^ was used to perform non-strand specific read counting for genes over exon features based on the Gencode V19 gene model (without excluding read duplicates). When both read pairs aligned uniquely (indicated by a STAR alignment quality score of 255) they were used towards gene reads counts. For total library abundance calculations, during TPM and FPKM expression values estimation, genes on chromosomes X, Y, MT and rRNA and tRNA were omitted as they can introduce library size estimation biases.

Hierarchical consensus clustering was applied using the cola package (version 1.5.6) with “MAD” as top-value method and “kmeans” as partitioning method. Classification on CNS samples was applied using cola with “ATC” as top-value method and “skmeans” as partitioning method. All other parameters took default values^77^.

#### RNA dilution experiment

To further investigate the impact of brain tissue contamination in unsupervised clustering analysis of gene expression data on PCNSL, we performed a serial dilution experiment with total RNA from a PCNSL sample considered “pure” (LS-027, estimated tumor cell content > 80%) and a normal brain tissue control (CTRL). Total RNA from LS-027 was mixed with CTRL RNA with increasing concentrations (0%, 20%, 40%, 60%, 80%) and sequenced. The z-score transformed TPM expression levels for PCNSL group 1 and group 2 signature genes for the serially diluted H050-0027 sample was compared against the cohort and individually using clustering analysis.

#### Differential expression analysis to identify signature genes

Differential expression (DE) of genes was analyzed using DESeq2 (version 1.14.1) with default settings using raw read counts from featureCounts. Genes without any count in all samples were excluded from the analysis.

## Data availability

The raw sequencing data of the 51 CNSL samples has been deposited at the European Genome-Phenome Archive under the accession number EGAS00001005339. Accession to the ICGC MMML-Seq data is available via the data accession committee of the ICGC (see: www.ICGC.org, Huebschmann *et al*^*30*^). All somatic mutation calls and integrated mutations tables on which the analysis was performed can be downloaded from Zenodo (https://doi.org/10.5281/zenodo.5106351)

### Validation of whole-genome and RNA sequencing results

#### Sanger sequencing

Bidirectional Sanger sequencing (bSS) was performed i) to validate WGS results in the PCNSL/SCNSL study cohort if sufficient DNA quantity was available (total: n=35; PCNSL: n = 26; PCNSL-M: n = 1; SCNSL: n = 2; SCNSL: n = 3; EBV+: n = 2), and ii) to identify mutations of recurrently mutated candidates in a larger set of additional PCNSL/SCNSL FFPE samples (FFPE extension cohort). The following genes were analyzed: *MYD88, KMT2D (MLL2), HLA-B, SETD1B, HIST1H1E, HLA-C, CD79B, BTG1, MYC, TP53, TERT, GRHPR, TBL1XR1, DST, PRDM15, OBSCN, FAT4, GRP98*. BSS was performed as previously described^78^. The PCR primers for the genomic regions of interest are displayed in **Supplementary table 3**. Sequencing was performed at Eurofins Genomics, Ebersberg, Germany. 142/147 (97%) of the selected variants (allele frequency above 10%) identified by WGS were confirmed. The results are displayed in **Supplementary tables 4 and 5**.

#### Formaldehyde-fixed paraffin-embedded (FFPE) CNSL extension cohort

Candidate genes were validated in 31 additional FFPE specimens of PCNSL (n = 19), SCNSL (n = 9), and n = 3 EBV positive cases. The DNA was extracted using the QIAamp DNA FFPE Tissue Kit (Qiagen, Hilden, Germany) according to the manufacturer’s instructions. The following, recurrently mutated genes (exons) were investigated: *PIM1* (exons 1-4), *MYD88* (exon 5), *OSBPL10* (exon 1), *GPHPR* (exons 2, 4, 5), *TBL1XR1* (exons 7, 8, 10, 12, 14), *KMT2D* (exons 32, 34, 38, 48, 50), *KLHL14* (exon 2), *HLA-B* (exons 2, 3), *PRDM15* (exons 9-12), *GPR98* (exons 65, 70, 81), *DST* (exons 13, 14, 23, 24, 36), *OBSCN* (exons 32, 63, 64, 85), *FAT4* (exons 1, 9, 17), *HIST1H3D* (exon 2), *HIST1H1E* (exon 1), *TERT* (promoter region). The results are displayed in **Supplementary table 5**.

#### Fluorescence in situ hybridization (FISH)

FISH analysis was performed as previously described^79^. Briefly, 4-μm FFPE sections were deparaffinized, dehydrated and incubated in pre-treatment solution (Dako, Denmark) for 10 min at 95– 99 °C. Samples were treated with pepsin solution for 3 - 6 min at 37 °C, washed, dehydrated, air dried and incubated with the respective DNA probe: *FHIT*: ZytoLight SPEC FHIT/CEN 3 Dual Color Probe, Zytovision, Germany; *CDKN2A* (9p21.3): Orange, Biocare Medical, USA; Vysis CEP 9 SpectrumGreen Probe (Abbott), The Netherlands). The sections were sealed, denatured in humidified atmosphere at 82 °C for 5 min and then incubated overnight at 45 °C to achieve hybridization. After post-hybridization washing, slides were counterstained with 4′6-diamidino-2-phenylindole (DAPI) and analyzed using an automated scanning system (Duet, BioView Ltd. Rehovot, Israel; **Supplementary figure 1 C**).

#### Real-time quantitative PCR (RT-qPCR)

We performed SYBR Green quantitative real-time PCR (qPCR) measuring six amplicons covering the *CDKN2A/B* gene (**Supplementary figure 1 D**) as well as five amplicons covering the *FHIT, NOTCH4, SPIB*, and *MIR650* gene. The primer sequences are annotated in the **Supplementary table 3**. qPCR analysis was performed on ABI Prism 7900HT Sequence Detection System (Applied Biosystems, Foster City, CA, USA).

#### Immunohistochemical (IHC) procedures

Immunohistochemical stainings were performed on a Benchmark XT autostainer (Ventana Medical Systems, Tuscon, AZ, USA) with standard antigen retrieval methods (CC1 buffer, pH8.0, Ventana Medical Systems, Tuscon, AZ, USA) using 7-μm-thick frozen or 4-μm-thick FFPE tissue sections. The primary antibodies used are listed in **Supplementary table 3**. The iVIEW DAB Detection Kit (Ventana Medical Systems, Tuscon, AZ, USA) was used according to the manufacturer’s instructions. Sections were counterstained with hematoxylin, dehydrated in graded alcohol and xylene, mounted and coverslipped. IHC stained sections were evaluated by two skilled neuropathologists with concurrence. The DLBCL subtypes of GCB and non-GCB were categorized using CD10, BCL6, and MUM1 according to the Hans algorithm^8^.

## Results

### Study cohort

We enrolled CNSL samples from 51 adults diagnosed with PCNSL or SCNSL. According to the site of manifestation, the following subgroups were defined: PCNSL within the brain parenchyma (PCNSL; n = 39), PCNSL with meningeal manifestation (PCNSL-M; n = 3), SCNSL within the brain parenchyma (SCNSL; n = 3), SCNSL with meningeal manifestation (SCNSL-M; n = 3) and EBV-positive lymphomas (EBV+; n = 3). Median age at diagnosis was 66 (range 40-82 years). The female:male ratio was 1.3:1. Follow up data was available for 44 patients. The follow up time ranged from 1 to 104 months with a median survival of 15.0 months (**Supplementary figure 1 E**). The detailed study cohort information and *s*ubgroup specific demographics are given in **Figures 1 A, B** and **Supplementary table 1**. Patient samples were histologically and immunohistochemically classified according to the WHO criteria^2,7,54,55^, and further stratified according to the Hans^8^ algorithm into non-GCB (n=37) and GCB subgroup (n=5, **Figure 1 C, Supplementary table 1**). For nine samples, the tissue was not sufficient for non-GCB/GCB characterization. Furthermore, we integrated data from the ICGC MMML-Seq cohort (www.icgc.org) for comparison of WGS and transcriptome data from systemic DLBCL, FL, naïve B-cells and GC B-cells^30,51,52^

#### Mutational landscape of central nervous system lymphoma (CNSL)

Whole-genome sequencing data of 38 CNSL (30 PCNSL, 1 PCNSL-M, 2 SCNSL, 3 SCNSL-M, and 2 EBV+ samples, **Figure 1 B**) was obtained with a median coverage of 66 (range 31-100) for tumors and 53 (range 27-85) for matched germline controls. We identified a median of 19577 (range: 1987-48280; median of the 30 PCNSL: 22199 (range 9185-48280)) total SNVs, of which a median of 6236 (range: 686-16731; PCNSL: 7058 (range: 2850-16731)) were intronic, a median of 10384 (range: 983-24033; PCNSL: 11797 (range: 5063-24033) were intergenic, and a median of 179 (range: 47-436), PCNSL: 202 (range: 100-436) were nonsynonymous exonic variants (1% of all SNVs). The number of detected SNVs was significantly higher in PCNSL compared to systemic DLBCL (*p* = 0.018; median SNVs: 18434 (range: 2,771-98,890)) or FL (*p* = 1.2 x10-11; median SNVs: 6049 range: 2139-19751), though different read-depths confound this comparison (median depth DLBCL (tumor): 32 (range: 27-38), (germline controls): 31 (range: 24-37); median depth FL (tumor): 32 (range: 27-36), (germline controls): 31 (range: 24-45)). Furthermore, we identified a median of 2958 (range: 711-9430; PCNSL: 3083 (range: 941-9430)) indels per CNSL sample, of which the majority was intergenic (1648 (range: 403-5218), PCNSL: 1723 (range: 517-5218). The mean number of variants (SNVs and indels) in non-coding RNA genes per patient was 2855 (range: 551-6913), PCNSL: 3201 (range: 1220-6913). Selected variants were verified using Sanger sequencing (**see Material and Methods section**).

The CNSL cohort presented a median of 188 (PCNSL: 191) SVs (range 24-517 (PCNSL: 47-517), inversions: 40 (PCNSL: 37), deletions: 99 (PCNSL: 107), duplications: 24 (PCNSL: 22), translocations: 20 (PCNSL: 21)). We also investigated chromosome level CNVs (based on 30% or more of a chromosome being amplified or deleted) and found a median of 9 CNVs (median 2 cnLOH (PCNSL: 2), median 4 gains (PCNSL: 4), and median 3 losses (PCNSL: 3)). The detailed mutational statistics (CNVs, indels, SNVs, and SVs) of the CNSL, DLBCL, and FL samples are displayed in **Supplementary table 6**.

#### PCNSL represent MCD genetic subtype of DLBCLs

Recent exome studies described the existence of different genetic subtypes of DLBCL, which show activation of distinct signaling pathways and different clinical outcomes^21,26,27^. We used the LymphGen algorithm described by Wright *et al*^27^ to classify our samples according to these genetic subtypes based on the obtained WGS data. The results of the CNSL cohort are displayed in **Figure 1 D**. In line with previous results^13,16,26,27^, the majority of PCNSL samples were classified as MCD (based on the co-occurrence of *MYD88* L265P and *CD79B* mutations, 67%, 20/30). One sample was each assigned to BN2 (*BCL6* fusions and *NOTCH2* mutations, 3%, 1/30) and ST2 (*SGK1* and *TET2* mutated, 3%, 1/30), seven samples were non-subtyped cases (“other”, 23%, 7/30), and one sample was equally assigned to both groups BN2/MCD (3%, 1/30; **Supplementary table 1**).

#### Driver mutations in CNSL

We first identified the genes recurrently mutated in CNSL (**Figure 2 A**) and used Metascape^80^ for further pathway and process enrichment analysis. The top three level enriched terms were ‘Regulation of hemopoiesis’, ‘Chromatin organization involved in negative regulation of transcription’, and ‘Cytokine signaling in immune system’ ((hypergeometric test, FDR 8.91×10-9, 1.04×10-4, 1.17×10-4, respectively; **Figure 2 B**). The enrichment analysis in TRRUST revealed ‘Regulated by: STAT3’ as the most significant term (hypergeometric test, FDR 3.98×10-7; **Figure 2 C**). *STAT3* has been associated with intracranial spreading and poor survival in PCNSL^81,82^, and reports of STAT3 inhibition via small molecules achieve complete tumor regression in vivo for lymphoma cell lines^83^.

**Figure 2:**
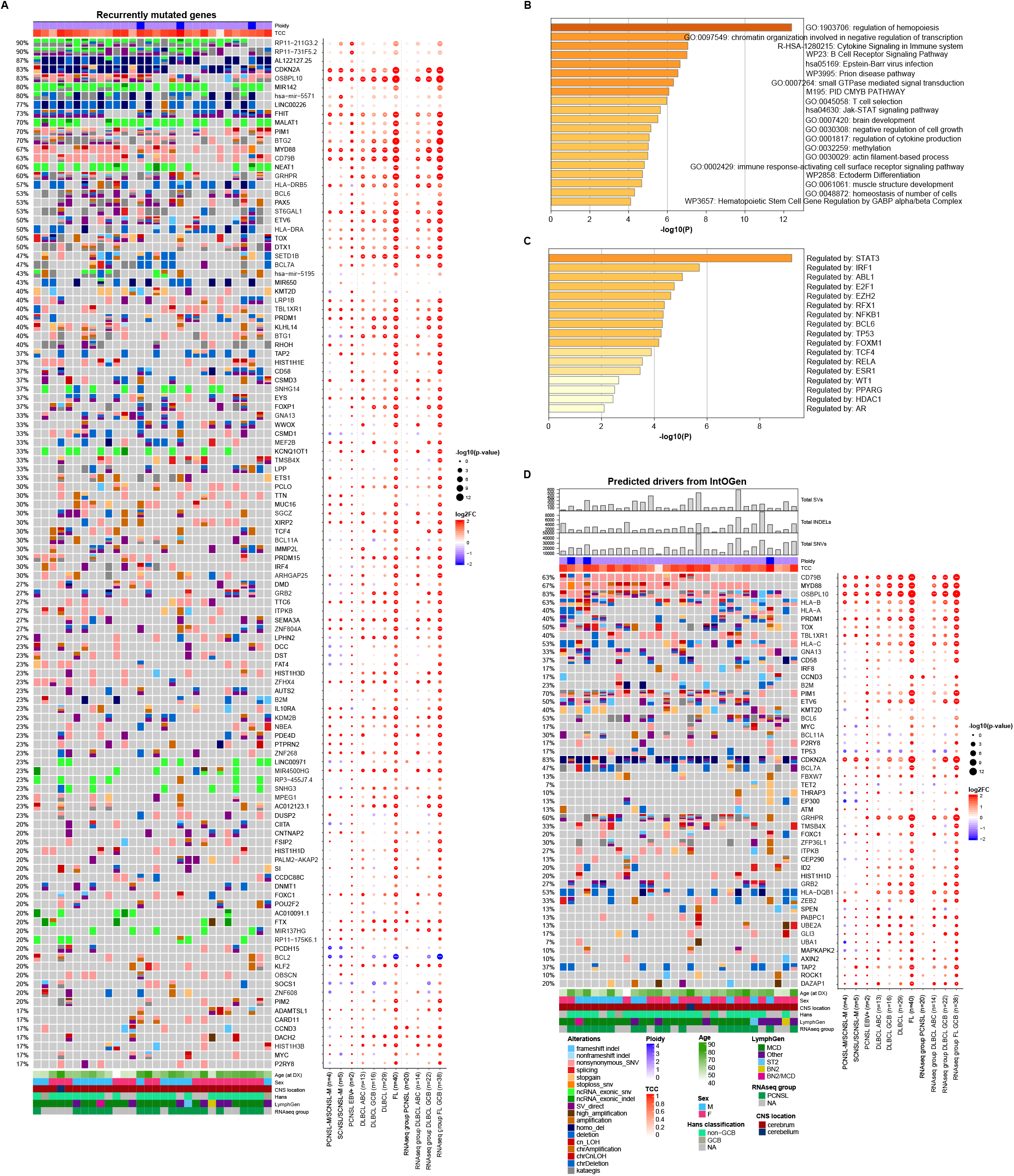
Recurrent coding mutations and genomic drivers in PCNSL. Oncoprints of recurrently mutated genes, excluding IG, chromosome Y genes (**A**). Mutated genes are listed from top to bottom depending on their alteration frequency. The corresponding dot plot reflects the log2 fold change and significance of alteration frequencies in the other subcohorts and RNAseq subgroups compared to PCNSL. The size of the dots demonstrate the significance according to a 2-tailed Fisher’
ss exact test. The recurrently mutated genes (listed in the oncoprint in A) were analysed by Metascape^80^ to identify pathway and process enrichment (**B**) and transcriptional regulatory networks (TRRUST, (**C**). Oncoprints of driver genes in PCNSL (**D**). The top panel of the oncoprint shows the total numbers of structural variants (SVs), small insertions/deletions (INDELs), single nucleotide variants (SNVs), estimated ploidy, and genomic tumor purity. Mutated genes are rankied by IntOGen. The corresponding dot plot reflects the log2 fold change and significance of alteration frequencies in the other subcohorts and RNAseq subgroups compared to PCNSL.

As *STAT3* is not highly mutated or hit by SVs or CNVs, its activation seems - in line with previous reports - induced by extrinsic factors such as infiltrating macrophages/microglial cells^84^, or intrinsic factors such as activation downstream of MYD88^85^.

Next, we used IntOGen and MutSigCV to discover putative driver mutations in the PCNSL WGS sub-cohort (**Figure 2 D, Supplementary Table 7**). We identified a total of 50 mutated driver genes, of which 21 were previously known drivers. Many of the predicted drivers were associated with MCD enriched genes, including *MYD88* (67%), *CD79B* (63%), *OSBPL10* (83%), *HLA-A/B/C* (40%/63%/53%), *PRDM1* (40%), *TOX* (50%), *TBL1XR1* (40%), *CD58* (37%), *PIM1* (70%), *ETV6* (50%), *BCL11A* (30%), *CDKN2A* (83%), *GRHPR* (60%), *FOXC1* (20%), *and DAZAP1* (20%). These driver genes were significantly enriched for genes containing the *BCL6* binding motif (TRANSFAC and JASPAR PWMs, Enrichr enrichment test, adjusted *p* = 0.03193).

Concerning *MYD88*, we only detected the classical pathogenic hotspot L265P mutation, which was validated by Sanger sequencing in all PCNSL samples investigated (100%, n = 26, **Supplementary tables 4 and 5**). Notably, *MYD88* mutation rates in the extension FFPE cohort were 78.9% in PCNSL and 55.6% in SCNSL. None of the five EBV-positive cases investigated harbored oncogenic *MYD88* L265P mutations, which is in line with previous findings^21^. Mutations in *TBL1XR1* also modulating TLR/MYD88 signaling^21^ were identified in 40% of PCNSL (**Figures 2 A, B**). We investigated mutual exclusivity and co-occurrence patterns for *MYD88* among the driver genes that affect at least five patients using Fisher and CoMET test. We observed mutual exclusivity between alterations in *MYD88* and the NOTCH signaling inhibitor *SPEN*^26^ (Fisher test, *p* = 0.0009, FDR = 0.033). In line with previous reports on ABC-DLBCLs^86,87^, we found coexisting alterations in *MYD88* and *CD79B*. Nevertheless, this co-occurrence was not significant (Fisher test, *p* = 0.16, FDR = 1.0). However *MYD88* was most significantly co-occurring with *TBL1XR1* (Fisher test, *p* = 0.04), both activating the NFkB signaling pathway^22^. Although this was not significant after correction for multiple testing (FDR = 1, **Supplementary table 8**).

Compared to the MCD driver genes identified in the series presented by Wright *et al*^*27*^, our PCNSL series exhibited a higher proportion of samples with mutations in *PABPC1* (10% vs 0%), *P2RY8* (13% vs 1.2%), *ITPKB* (23% vs 2.5%), *GNA13* (20% vs 5.1%), and *B2M* (13.3% vs 2.8%). Furthermore, predicted driver genes in our PCNSL series included genes enriched in all other LymphGen classes: BN2 (*CCND3, BCL6, HIST1H1D, SPEN, PABPC1*, and *UBE2A*), EZB (*GNA13, IRF8, BCL7A, KM2TD, EP300*), ST2 (*P2RY8, TET2, ZFP36L1, and ITPKB*) and A53 (*B2M* and *TP53*).

While the majority of identified drivers were reported by Wright *et al*, a number were not, including *FBXW7, ATM, TMSB4X*, THRAP3, *ID2, GRB2, ZEB2, GLI3, UBA1, MAPKAPK2, AXIN2, TAP2, ROCK1, CEP290*, and *HLA-DQB1*. These were previously recognized as general DLBCL drivers by Reddy *et al*^88^ and/or Chapuy *et al*^25^. *ZEB2* was additionally identified as a genetic alteration associated either with the ABC subgroup^88^ or the DLBCL C1 cluster^25^ (**Supplementary table 7**).

#### Recurrent somatic alterations in non-protein-coding genes

We identified recurrent aberrations in several non-protein coding genes (**Figure 3 A**) which mainly affected short and long non-coding RNA (ncRNA) such as the aberrant SHM (aSHM) target *MIR142* (80%; **Figures 3 A, B**) as well as *MALAT1* (70%) and *NEAT1* (60%), both located 53 kb apart on 11q13.1. The mechanistic roles of many ncRNAs are poorly understood because their exact function is difficult to assess. However, the lncRNAs *NEAT1* (nuclear enriched abundant transcript 1) and *MALAT1* (metastasis-associated lung adenocarcinoma transcript 1) are well known to play essential roles in the development and progression of various cancers by influencing gene expression by alternative splicing and epigenetic modification of regulatory elements^89-91^. Further aberrations in lncRNAs affected *KCNQ1OT1* (33%) and *SNHG3* (23%), both reported to have oncogenic functions in multiple cancers^92,93^ as well as *SNHG14* (37%), promoting immune evasion in DLBCL^94^.

**Figure 3:**
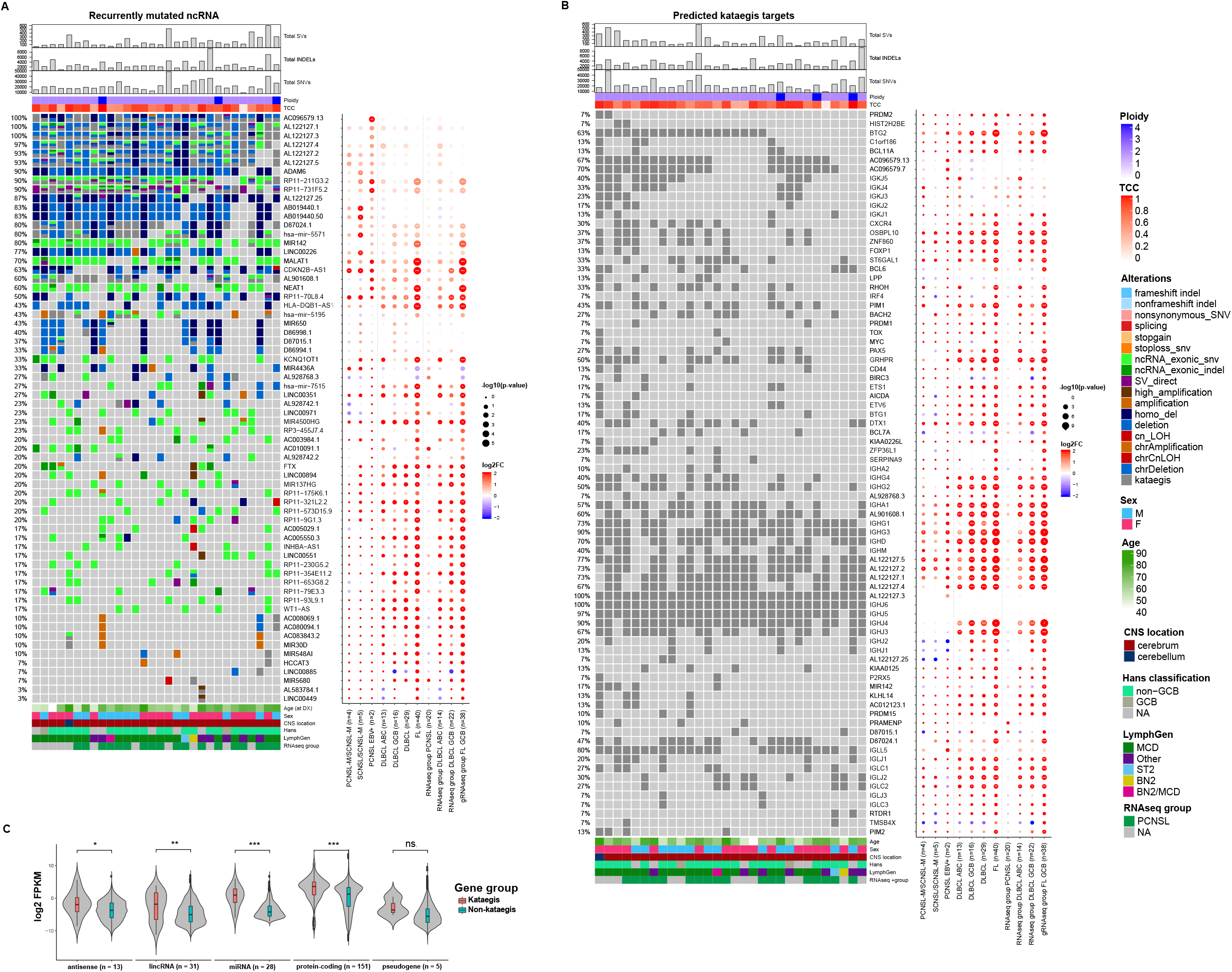
Recurrent non-coding RNA mutations and Kataegis events in PCNSL. Oncoprint of recurrent non-protein-coding genes in PCNSL (**A**). Shown are the total numbers of structural variants (SVs), small insertions/deletions (INDELs), single nucleotide variants (SNVs), estimated ploidy and tumor purity (top panel). Mutated genes are listed from top to bottom depending on their alteration frequency. Oncoprint of recurrent kataegis events in PCNSL (**B**). Mutated genes are listed from top to bottom depending on chromosome location. The corresponding dot plot reflects the log2 fold change and significance of alteration frequencies in the other subcohorts and RNAseq subgroups compared to PCNSL. The size of the dots demonstrate the significance according to a 2-tailed Fisher’
ss exact test. The violin plot (**C**) shows that the RNA expression of genes with kataegis loci compared to those without were significantly higher for antisense, long non-coding RNA, miRNA and protein coding genes (Wilcoxon rank sum test, *p* < 0.05).

#### Kataegis shapes the mutational repertoire of PCNSL

Kataegis is a pattern of mutational hotspots that has been associated with a number of cancers^68^, and is a frequent consequence of AID activity in lymphomas^95^. Many of the recurrently mutated genes in PCNSL were dominated by alterations that are located in these highly mutated hotspots^11^, of which several have previously been described as targets of aSHM, such as *OSBPL10, PIM1, BTG2*, and *PAX5* (**Figure 3 B**)^21,45,96^. Of the 50 identified protein-coding driver genes and the top 50 mutated ncRNA in PCNSL, 15 and 21 were targeted by kataegis, respectively (**Figures 2 A, B, 3 A, B, additional supplement (**https://doi.org/10.5281/zenodo.5106351). Consistent with previous reports^97^, expression of miRNA, lncRNA, antisense RNA, and protein coding genes with kataegis loci were expressed significantly higher than those without (Wilcoxon rank sum test, *p* < 0.05; **Figure 3 C**). This implicates that either aSHM preferentially targets highly expressed genes, or that aSHM may cause hyperactivation of these genes. Interestingly, the largest difference in RNA expression was observed in miRNA genes, again highlighting the importance of the non-coding alterations in PCNSL. This observation was consistent for subgroups, including systemic DLBCL (**Supplementary Figure 2**).

Physiologically, SHM is the process of introducing mutations in the antibody genes to alter the antigen-binding site, increasing the immunglobulin (IG) diversity^98^. Kataegis events were at *IGH* (100%), *IGL* (100%) and *IGK* (70%) loci but were also found outside IG loci, targeting *BTG2* (63%), *GRHPR* (50%), *PIM1* (43%), *DTX1* (40%), *OSBPL10* (37%), *ZNF860* (37%), *BCL6 (33%), RHOH* (33%), *CXCR4* (30%), *BACH2 (27%)*, and *PAX5* (27%; **Figure 3 B**). The recurrently targeted genes in PCNSL mostly overlapped with those targeted in ABC-DLBCLs. However, samples with mutational hotspots in *BTG2, GRHPR, OSBPL10* and *ZNF860* were significantly more frequent in PCNSL (in 18, 15, 11 and 11 of 30 samples, respectively) compared to ABC-like DLBCL (in 2, 0, 0 and 0 of 13 samples, *p* = 0.009, 0.001, 0.019 and 0.019, Fisher’
ss exact test, respectively). Taking all non-immunoglobulin genes that overlapped a mutational hotspot in at least one PCNSL sample (242 genes, **Supplementary table 9**), we found the BCR signaling pathway to be most significantly enriched (Enrichr enrichment test, adjusted *p* = 0.0046). Taken together, kataegis and aSHM play a decisive role in shaping the mutational repertoire of PCNSL and are associated with functional pathways in PCNSL pathogenesis.

While patterns of aSHM and kataegis were similar between CNSL and systemic DLBCL subtypes, we identified that EBV+ CNSL cases did not share many of the recurrent mutational hotspots apart from IGH and the *HLA−DRB locus*. (**Figure 3 B, Supplementary figures 3 A, B, Supplementary table 5**).

#### Recurrent copy number alterations (CNAs)

Compared to systemic ABC-DLBCL^52,30,99^, PCNSL demonstrated significantly more CN losses in 6p21 (*HLA-D locus*, **Figure 4 A, B, Supplementary table 10**) as well as recurrent losses in 9p21 (*MTAP, CDKN2A/B*) and 19p13 (*CDKN2D*). The loss of the *HLA-D* locus that encode for MHC class II molecules lead to reduced immune surveillance and poor survival in DLBCL^100^. *CDKN2A* is an established tumor suppressor gene with roles in angiogenesis, cell death, invasiveness, and growth suppression^101-103^. Additionally, we found deletions on chromosomes 1p13 and 3q13, affecting genes such as *CD58* and *CD80*, both candidates reported to lead to immune evasion^104^. Further CN losses were detected on chromosomes 8q12 (*TOX*), 12p13 (*ETV6*), and 15q21 (*B2M*) as well as 3p14, affecting the fragile site tumor suppressor gene, fragile histidine triad (*FHIT*). *TOX* deletions have been previously described by array-based imbalance profiling^105^. *TOX* is required for the development of various T-cell subsets and was described as putative tumor suppressor in MCD DLBCL^26^. *TOX* downregulation has been associated with poor prognosis in different cancers^106^ and is a predictor for anti-PD1 response^107^. Significant CN gains in PCNSL mapped to 2q37 and 18q21 affecting *DIS3L2* and *MALT. DIS3L2* encodes for an exoribonuclease that is responsible for Perlman syndrome^108^ and was recently described to promote HCC tumour progression by upregulating production of the oncogenic isoform of *RAC1, RAC1B*^109^. *MALT* is a regulator of NFkB signaling and potential therapeutic target in B-cell lymphoma^110^.

**Figure 4:**
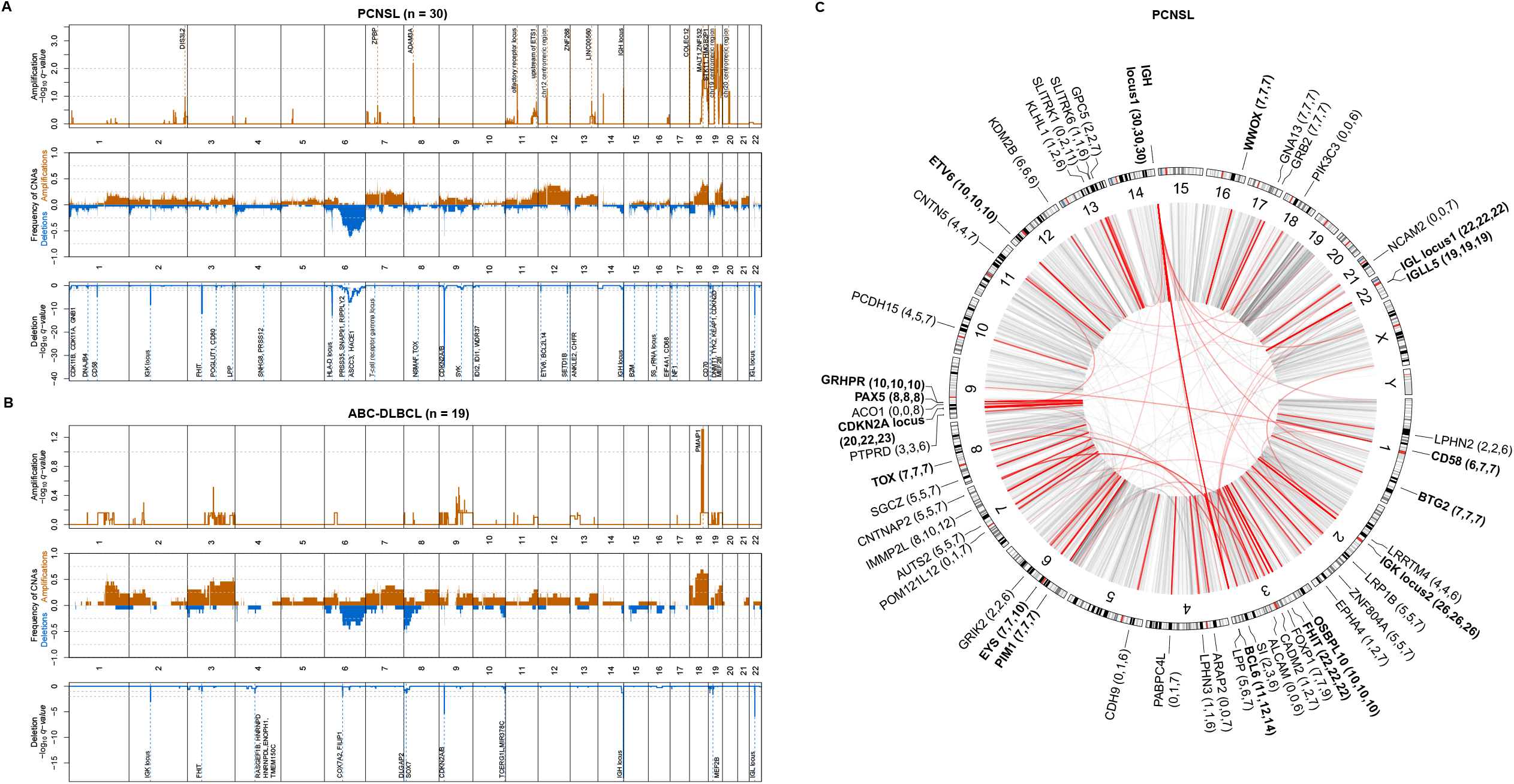
Genomic structural variation in PCNSL. Recurrent somatic copy number aberrations in PCNSL (**A**) and ABC-DLBCL (**B**). Relative prevalence of somatic copy number aberrations in tumour samples (middle panel), showing presence of at least one copy number gain (orange bars), copy number loss (blue bars), as a proportion of analysed samples. Significantly (q-value < 0.25) amplified and deleted regions and candidate genes are shown (upper and lower panel). Circular visualization of genome rearrangements in PCNSL (**C**). The panels (from outside going inwards) represent recurrence per gene, chromosome ideograms, and chromosome numbering. Next to gene names, the number of SV breakpoints that lie direct on the gene, within 100 kbp of the genes, and closest to that gene are reported. Inter-chromosomal translocations are rendered with black (all) and red (highlight 6+) arcs in the centre of the display.

#### Recurrent structural variations (SVs)

We defined SVs as genomic breakpoints, which can correspond the borders of amplifications and deletion, but also balanced translocations and inversions. PCNSL showed a median of 192 SVs (range 24-517, **Supplementary table 11**). Immunoglobulin gene rearrangements were found in all PCNSL cases and affected the *IGH* (100%), *IGL* (67%) and *IGK* (93%) loci (**Figure 4 C**). Furthermore, in PCNSL direct SVs affected *FHIT* (77%), *CDKN2A*/*B* (67%), *BCL6* (37%), *OSBPL10* (33%), *ETV6* (33%), *PAX5* (27%), *PIM1* (23%), *TOX* (23%), *BTG*2 (23%), *WWOX* (23%), as well as *CD58* (20%) **Figure 4 C**). *WWOX* and *FHIT* represent common fragile site (CFS) and have been classified as tumor suppressor genes in DLBCL^111,112^.

Recent studies have shown that translocation can act as enhancer hijacking even when the events is several hundred thousand base-pairs away from target genes^113^. To investigate this, we also annotated SV breakpoints to genes within 100 kbp and also to the closest genes. We found a number of genes involved in G protein-coupled receptor signaling (*ARAP2, LPHN2, LPHN3, EPHA4, ADGRL2*, and *GPC5*) consistent with observations in pan-cancer studies^114^. A number of other genes exhibited at least three times as many distal translocations (while still being the closest gene) than directly on the gene, including *PIK3C3, EPHA4, SI, ALCAM, NCAM2, CADM2, CDH9, PABPC4L, GRIK2, POM121L12, ACO1, KLHL1, SLITRK1*, and *SLITRK6*. Hyperactivation of PI3K signaling is one of the most common events in human cancers, and PIK3C3 has been shown to promote cell proliferation^115^ and autophagy^116^, and its inhibition has shown therapeutic benefit in bladder, HCC and colon cancer^117-119^. *EPHA4* has been described to promote cell proliferation and migration^120,121^ and was associated with tumour aggressiveness and poor patient survival in human breast and rectal cancer^122,123^. Inhibition of EphA4 has been shown to overcome intrinsic resistance to chemotherapy^124^. Many of these other potential enhancer-hijacking targets do not have well established roles in cancer pathogenesis, however we did notice a number of genes involved in cell adhesion (A*LCAM, NCAM2, CADM2*, and *CDH9*) and 2 SLIT and NTRK like family members (*SLITRK1, SLITRK6*).

#### Immunoglobulin translocations implicate distinct CNSL subtypes

IG translocations are established oncogenic drivers of many lymphatic neoplasms^125-127^. *IGH-BCL6* fusions are recurrent in PCNSL^20^, which mirrors observations of ABC-DLBCL^128^. *IGH-BCL2* fusions are more prominent in GCB-DLBCL^129^. We investigated the recurrent translocations (≥ 2 patients) in our cohort and identified five CNSL samples with *IGH-BCL6* translocations (**Figure 5 A, Supplementary figures 4 A-D**). We also identified three cases with *IGH-BCL2* translocations (**Figure 5 B, Supplementary figures 4 E, F**) one in each of SCNSL, SCNSL-M and PCNSL-M, further implicating that meningeal and secondary CNSL are distinct from intraparenchymal PCNSLs. Two PCNSL cases showed *IGL* and *IGH* translocations with breakpoints close to *CD274* (PD-L1; **Figure 5 C, Supplementary Figure 4 G**), which resulted in strong PD-L1 protein expression (**Supplementary Figure 4 H)** and therefore implicates a potential target for immunotherapy. All other, non-recurrent translocations are listed in **Supplementary table 12**.

**Figure 5:**
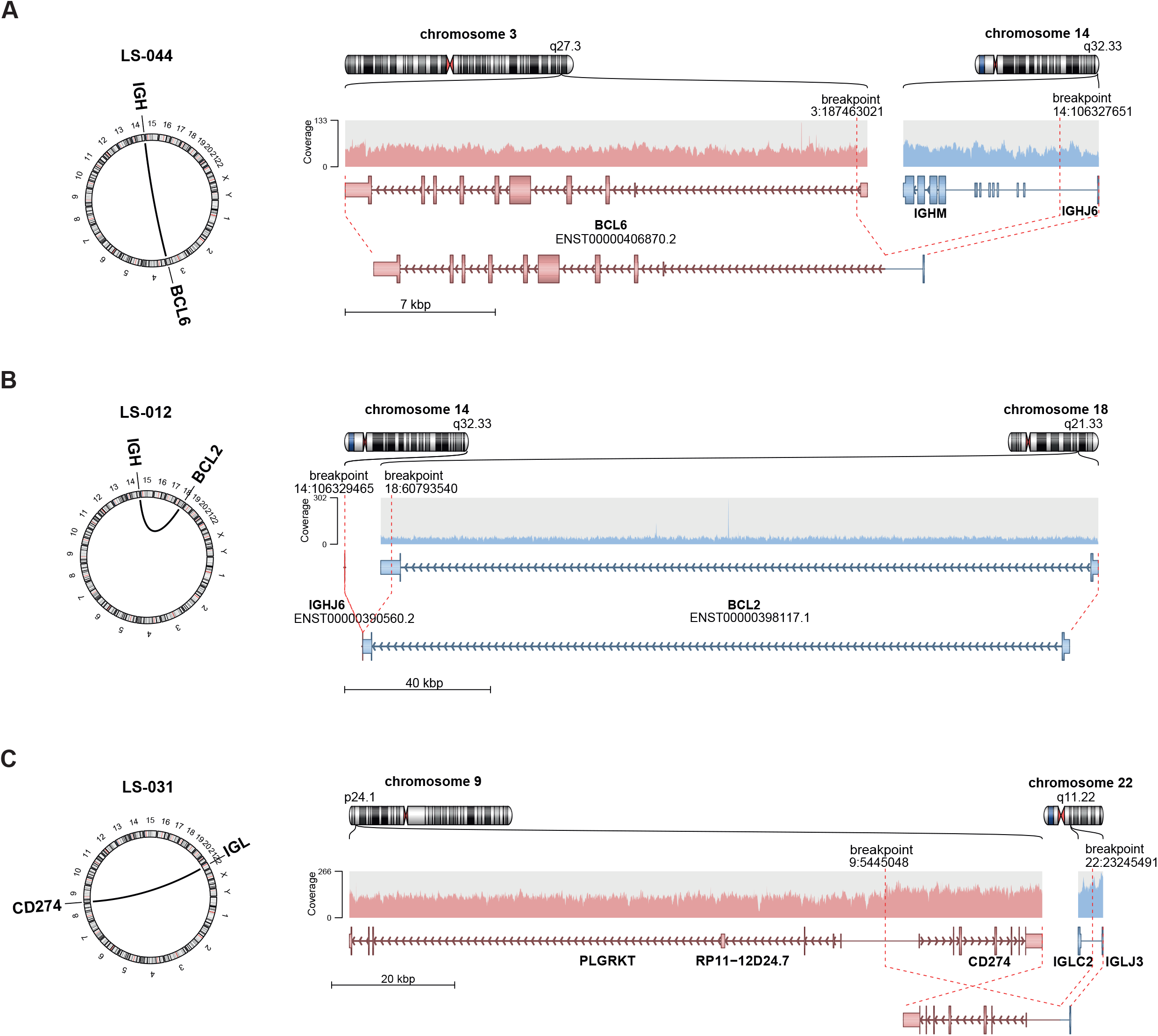
Immunoglobulin translocations in PCNSL. Schematic representation of the translocation breakpoints involving *BCL6* (PCNSL patient LS-044; **A**), *BCL2* (SCNSL-M patients LS-012; **B**), and *CD274* (PD-L1; **C**). The left panels show a circular plot displaying the location of the translocation partners. The right panel shows the original and reconstructed translocation events. Shown are (from top to bottom) chromosome ideograms, read overage of the sites, the gene models, and the reconstructed translocation partners.

The analyses of the IG breakpoints provided in all informative junctions evidence that these occurred due to illegitimate CSR or aberrant SHM, with the notable exception of the *IGH-BCL2* junctions, which were the consequence of an aberrant VDJ rearrangement (**Supplementary table 13**). Thus, all IG translocations in PCNSL are supposed to occur in the germinal center process rather than in a pre B-cell.

#### Mutational signatures in PCNSL

Mutational signatures were analyzed with regard to SNVs (single base substitutions, SBS) and indels (ID) of all tumor samples as defined by Alexandrov et al^70^ (**Figure 6**). For single base substitution signatures (SBS) we found mutational patterns that have been associated with spontaneous deamination of 5-methylcytosine (SBS1), defective activity of the AID/APOBEC family (SBS2), failure of double-strand DNA break repair by homologous recombination (SBS3), somatic hypermutation (SBS9), and damage by reactive oxygen species (SBS18). Additionally, the samples frequently revealed mutations caused by mutational signatures SBS5, SBS17b, and SBS40, which are of unknown etiology (**Figure 6 A**). The presence of SBS3, hallmark of defective DNA break repair by homologous recombination, and SBS40 may be therapeutically relevant as these indicate potential effectiveness of combination therapy with PARP inhibitors (e.g., Olaparib) alongside cytotoxic chemotherapy^130,131^. The three most prominent signatures in DLBCL, FL and CNSL were SBS9, SBS5, and SBS40 (**Figure 6 B**). Direct comparison of PCNSL and DLBCL revealed that signature SBS1, which correlates with DNA replication at mitosis (mitotic clock)^70^, was significantly enriched in PCNSL (*p* = 0.0027; **Figure 6 C, Supplementary figures 5 A-G)**.

**Figure 6:**
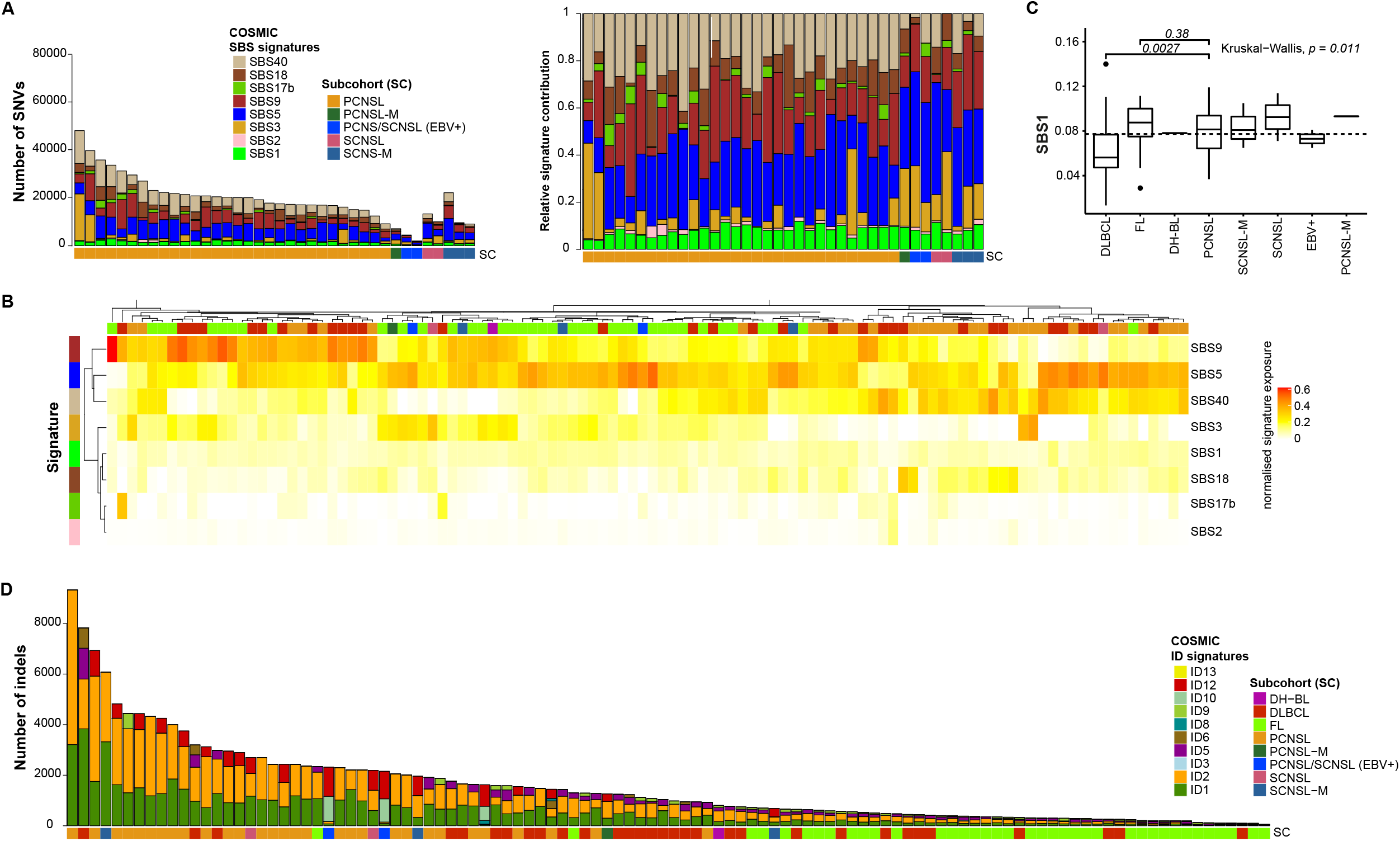
Mutational signatures in PCNSL. Single base substitution (SBS) signature contribution in PCNSL and SCNSL. Stacked bar plot are ordered by subgroup and then decreasing mutations load (**A**). The left bar plot shows the number of SNVs and the right shows the normalised signature exposures. Each colour corresponds to a mutations signature and the proportion of the colour reflects the number or proportion of SNVs explained by a certain mutational signature. The heatmap shows the clustering pattern of the SBS mutational signatures in all CNSL and peripheral lymphomas (**B**), which revealed groupings mutational signatures SBS5, SBS9, and SBS40. Pairwise comparison of PCNSL with DLBCL or FL (Mann-Whitney U test) revealed that signature SBS1 was significantly enriched in PCNSL compared to DLBCL (*p* = 0.027; (**C**)). Small insertion and deletion (ID) signatures in CNSL and peripheral lymphoma (**D**) demonstrate elevated numbers of indels and mutational patterns associated with slippage during DNA replication of the replicated DNA strand (ID1) and template DNA strand (ID2) in PCNSL.

Analysis of small insertion and deletion signatures (ID) revealed mutational patterns associated with slippage during DNA replication of the replicated DNA strand (ID1) and template DNA strand (ID2); both of these signatures appeared significantly (*p* < 1×10^−4^, Wilcoxon) more prominent in PCNSL compared to DLBCL and FL (**Figure 6 D**), though different read-depths may have influence this analysis.

Interestingly, only CNSL samples but not DLBCL or FL revealed mutations caused by mutational signature ID12 that is of unknown etiology and has been observed in prostate adenocarcinoma and soft tissue liposarcoma^70^.

### PCNSL RNA expression signatures are distinct from systemic DLBCL

The relative rarity of PCNSL and limited availability of fresh frozen tissue have thus far complicated the implementation of larger molecular studies needed for patient stratification. To unravel the molecular signature of PCNSL, we employed an unsupervised consensus clustering approach (using the cola tool^77^) to identify expression groupings between PCNSL samples and samples from the ICGC MMML-seq project (mainly consisting of non-GCB and GCB type DLBCLs, and FLs). This yielded the following major clusters: FL, PCNSL, GCB type DLBCL, ABC type DLBCL, non-tumorous GC-B-cells, and naïve B-cells (**Figure 7 A**). For each cluster, we identified signature gene sets that significantly correlated with the groupings. Interestingly, all meningeal PCNSL (PCNSL-M) and SCNSL-M grouped together with either GCB- or ABC-DLBCL, clearly indicating that these subtypes are molecularly and pathomechanistically distinct from intraparenchymal CSNL, which formed one separate cluster suggesting a distinct signature of CNS tropism. The ABC type DLBCL cluster was enriched for *MYD88* mutant samples, which were still distinct from *MYD88* mutant PCNSL at the gene expression level. To further exclude an impact of the potentially contaminating surrounding CNS tissue, we analyzed total RNA from normal brain controls (n = 2), of which one control was spiked with increasing concentrations of RNA from a pure PCNSL sample (0%, 20%, 40%, 60%, 80%). Then, we further stratified the PCNSL group by another round of consensus clustering using the two different classification methods (**Figure 7 B, Supplementary figures 6 A-C**), which both revealed two groups based on tumor purity.

**Figure 7:**
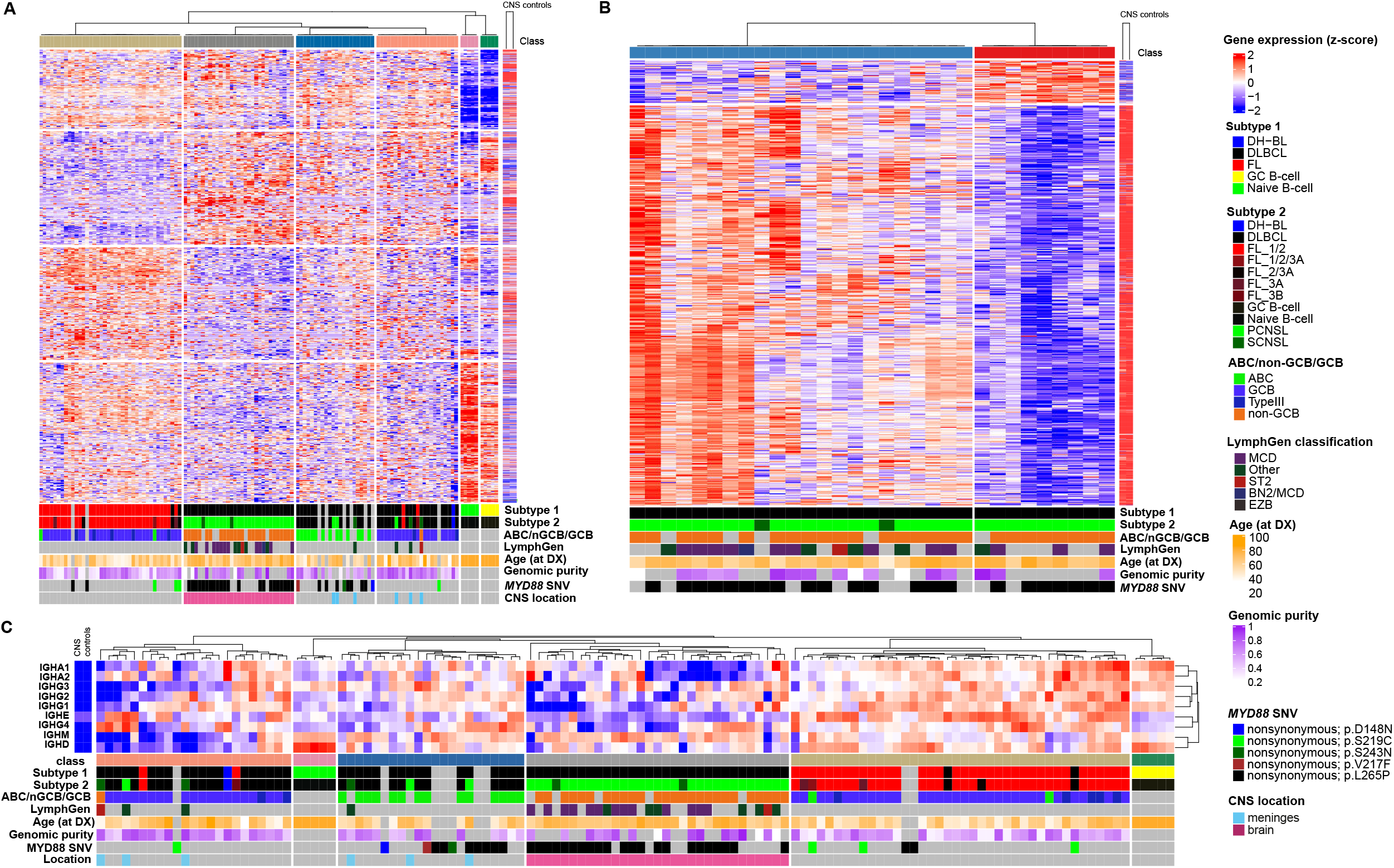
Transcriptomic signatures distinguish PCNSL from DLBCL. Global gene expression patterns clearly separate PCNSL from ABC-DLBCL, GCB-DLBCL, and FL (**A**). The heatmap shows unsupervised consensus clustering of gene expression data. All SCNSL-M and PCNSL-M cluster with non-CNS-DLBCL, distinct from intraparenchymal PCNSL. The ABC-DLBCL cluster is enriched for *MYD88* mutant samples, which are distinct from *MYD88* L265P mutant PCNSL. Samples of normal germinal center (GC) and naïve B-cells were included as controls. Furthermore, normal brain tissue (CNS controls) samples were added to analyse the impact of normal brain tissue contamination at the RNA level. Consensus clustering (skmeans) with intraparenchymal PCNSL samples and CNS controls revealed two groups (**B**). The first PCNSL expression group (PCNSL1, “pure”, right) consisted of samples with high tumor cell content, the second (PCNSL2, “impure”, left) contained mainly samples with a lower tumor cell content, which signatures correlated well with normal brain tissue expression. PCNSL can be distinguished from ABC-DLBCL, GCB-DLBCL and FL based on the expression of *IG* constant genes (**C**).

The first PCNSL expression group (PCNSL1, “pure”) consisted of samples with high tumor cell content (determined by whole-genome sequencing and histopathological analysis). Expression of its signature gene set did not show similarity to normal brain tissue expression. However, the second PCNSL expression group (PCNSL2, “impure”) contained mainly samples with lower tumor cell content, and expression of its signature gene set was indeed similar to normal brain tissue expression. We identified the PCNSL signature gene sets relative to ABC and GCB type DLBCLs and FLs, and removed potential background signatures from contaminating brain tissue (**Figure 7 B**). The marker genes in each group were identified based on differential gene expression analysis (**Supplementary table 14**). Among the marker genes for PCNSL were e.g. *LAPTM5*, a CD40 related gene expressed in malignant B-cell lymphoma^132^ and *ITGAE*, mediating cell adhesion, migration, and lymphocyte homing through interaction with E-cadherin^133^. We used Metascape^80^ for functional analysis of marker genes. Further pathway and process enrichment analysis revealed that pathways such as ‘ribonucleoprotein complex biogenesis, ‘mRNA processing’, ‘cell cycle’, ‘RNA modification’, ‘DNA conformation change’, and ‘DNA-templated transcription, initiation’ were enriched (**Supplementary Figure 6 D**). The top three level Gene Ontology biological processes included ‘cellular component organization or biogenesis’, ‘metabolic process’, and ‘localization’ (**Supplementary Figure 6 E**).

#### Expression of IGHM is characteristic for PCNSL

Additionally, we analyzed the expression of *IG* constant genes, which again revealed the same clusters as the unsupervised consensus clustering approach, demonstrating that PCNSL can be differentiated from DLBCL based on only the expression of *IG* constant genes. In contrast to DLBCL and FL, PCNSL show generally low expression of *IG* constant genes, but higher expression of *IGHM* (**Figure 7 C**).

#### *TERT* expression but not telomere content upregulated in PCNSL

Telomerase activity and telomerase reverse transcriptase (*TERT*) gene expression have been reported as prognostic factors in PCNSL patients^134^. We used TelomereHunter, a software for detailed characterization of telomere maintenance mechanisms^73^ to estimate the telomere content in a representative cohort of PCNSL, SCNSL, peripheral lymphoma, as well as non-tumorous naïve and GC-B-cells as control^51^. In approximately 1/3 of the samples, the purity-corrected telomere content was higher in the tumor than in the matched control (whole blood) (**Supplementary figure 7 A**). Nevertheless, telomere content (tumor/control log2 ratio) was not significantly different between the different histological, clinical and molecular subgroups (**Supplementary figure 7 B**). However, expression of the *TERT* gene, the main activity of the encoded protein is the elongation of telomeres, was significantly higher in GC B-cells^51^ and in PCNSL compared to ABC-DLBCL (**Figures 8 A, B**). This was consistent with observations when stratifying samples by RNA subgroups, where *TERT* expression was significantly higher in PCNSL compared to ABC-DLBCL, GCB-DLBCL and FL (**Figure 8 C**).

**Figure 8:**
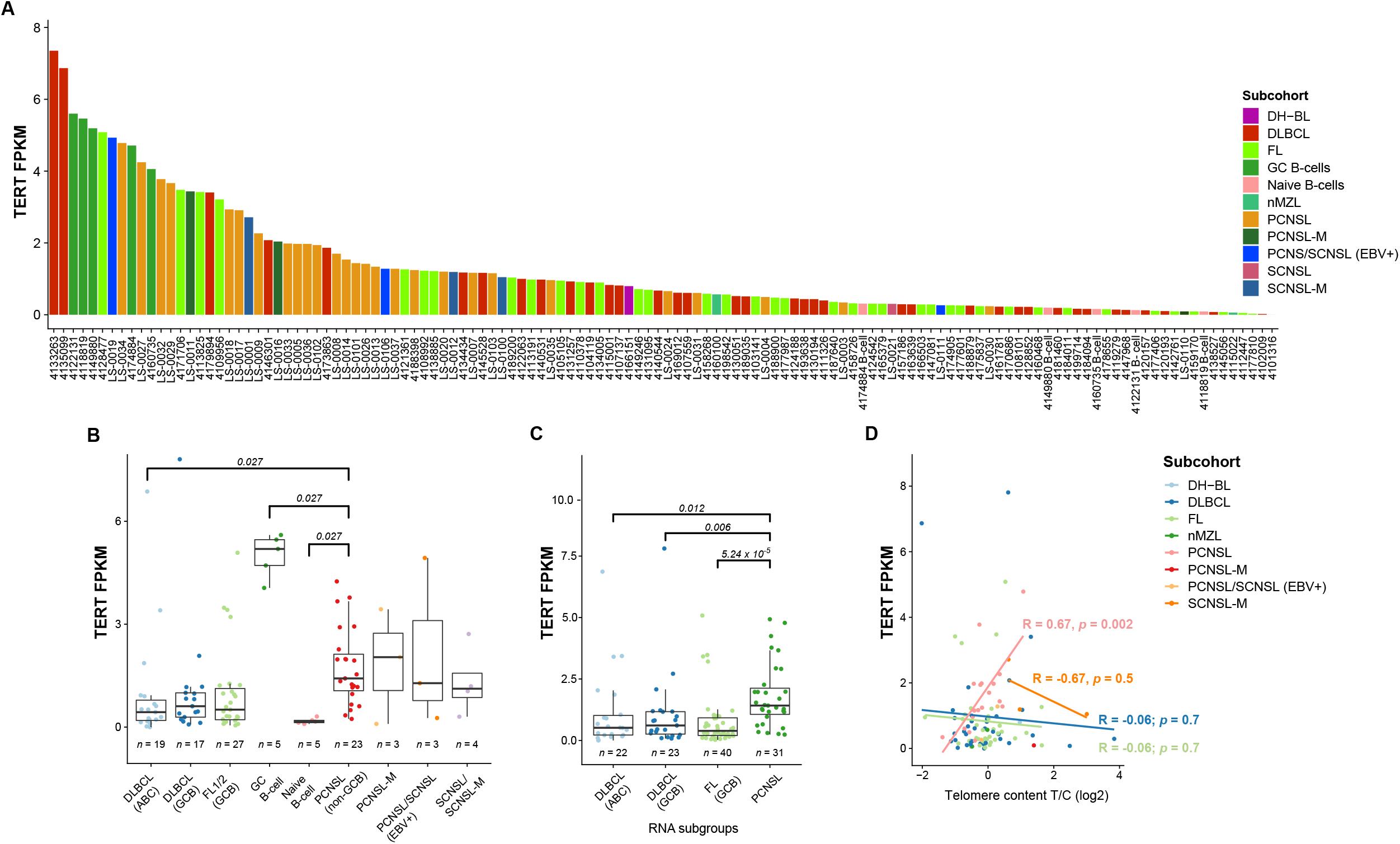
*TERT* expression correlates with telomere content in PCNSL. *TERT* expression in each sample of the CNSL and peripheral lymphoma cohort as well as in in the different subcohorts (**B**), and RNAseq groups (**C**). *TERT* expression was significantly higher in PCNSL compared to both ABC-DLBCL and GCB-DLBCL (using Holm-Bonferroni method for statistical testing). A high positive correlation between *TERT* expression and telomere content was only observed in PCNSL (**D**).

Interestingly, the higher *TERT* expression in PCNSL significantly correlated with normalized telomere content (Pearson’s R = 0.67, *p* = 0.0023; **Figure 8 D**). However, SCNSL-M, DLBCL and FL did not show such trends (R = -0.67, *p* = 0.53, R = -0.065, *p* = 0.70, and R = -0.061, *p* = 0.72, respectively). This suggests that *TERT* has an active role in combatting telomere degradation in PCNSL. Two well-known promoter hotspot mutations (−124C>T (C228T) and −146C>T (C250T)) have been described to increase *TERT* expression and cell-cycle progression^135,136^. These mutations have been found in several solid and hematological malignancies including different brain tumors and PCNSL^137-139^. Therefore, we next investigated the *TERT* promoter mutation status in our WGS (n=38) and FFPE extension cohort (n=31). Sanger sequencing of the *TERT* promoter region was performed i) for all WGS samples having only low coverage in the promoter sequence (below 40x, n = 6, **Supplementary figure 7 C**), ii) the FFPE extension cohort, and iii) three oligodendrogliomas, known to carry high frequency *TERT* promoter mutation^140^. We detected no *TERT* promoter mutations in 67 samples of PCNSL and SCNSL (**Supplementary table 5, Supplementary Figure 7 D**), while the well-known TERT rs2853669 polymorphism, which has been associated with increased cancer risk^141^, was identified in 40% (14/35 (8 PCNSL, 6 SCNSL)) of the patients in the extension FFPE cohort. The Sanger sequencing results of two samples were not conclusive.

## Discussion

Here we have performed a comprehensive analysis of recurrent protein coding and non-coding mutations, CNVs, structural variants, and driver mutations in the largest cohort of PCNSL to date and compared the genetic features to systemic DLBCL and -FL. The vast majority of PCNSLs are of non-GCB DLBCL subtype^142^ and share many genetic alterations with non-CNS ABC-DLBCL in the same signaling pathways. Previous studies made use of whole-exome sequencing^21,45^ which i) limits the investigation to protein-coding regions and ii) may not be ideal for understanding the patterns of mutational hotspots - e.g. attributed to AID induced somatic hypermutation in B-cell non-Hodgkin lymphomas^143^ - as well as the structural variation in genomes^144^. PCNSL showed significantly more SNVs and indels compared to systemic DLBCL, even in intronic and intergenic regions, also underlining the importance of non-protein coding aberrations in PCNSL pathogenesis. Many of the recurrent mutations in non-protein coding genes affected non-coding RNAs (ncRNAs), which are among other functions involved in epigenetic regulation of gene expression, cell differentiation, and development^89,145^.

In line with previous results^21,37,146^, we here demonstrate that PCNSL are defined by recurrent and often biallelic *CDKN2A/B* deletions, *MYD88* L265P mutations, and mutations that activate B-cell receptor (BCR) signaling, genetic hallmarks of the DLBCL subtype MCD/C5^26,27^. Furthermore, we found high frequencies of SVs affecting the *IGH, IGL* and *IGK* loci as well as losses of chromosome 6p affecting the *HLA* gene complex as a mechanism to escape recognition by cytotoxic T-cells^147^. *MYD88* L265P mutation and *CDKN2A/B* loss have been described as early mutational events in PCNSL^39^ and we confirmed both to be major drivers in PCNSL. While *TP53* alterations seem to play a minor role in PCNSL, the *CDKN2A/B* genes encode several proteins that regulate either the p53 (p19 ARF) or the RB1 (p16 INK4a) pathway^148,149^, underlining the relevance of the TP53 pathway in the context of PCNSL and cell cycle control.

The frequencies of *MYD88* mutations had varied between 38% and 94.4% in previous PCNSL studies^22,27,45,46,150^, which might reflect a selection bias among small study populations, given the rarity of PCNSL. This huge range could alternatively result also from an imprecise definition of PCNSL, which includes all malignant NHL within the brain, eyes, spinal cord or leptomeninges without systemic involvement. In contrast, we here defined PCNSL as only intraparenchymal CNS-DLBCL and found a high prevalence of the *MYD88* L265P variant in this cohort (WGS cohort, extension FFPE cohort; median: 73%). This is further supported by our robust classification of PCNSL by the RNA sequencing results, which demonstrated that the expression profiles of PCNSLs were distinct from PCNSL-M, SCNSL-M, and peripheral DLBCL without CNS manifestation, the latter three entities sharing similar profiles.

Moreover, somatic hypermutation has previously been described as having a pathogenic role in PCNSL development and that its extent was greater there than in systemic DLBCL^96^. In agreement with previous reports, we identified several aSHM targets including the proto-oncogenes *PIM1, PAX5, BTG2*, and *OSBPL10*^21,45,96^. Exploiting a whole-genome sequencing approach, we observe additional mutational hotspots indicative of aSHM also in other genes including *MIR142, FHIT, ETV6, BTG1, GRHPR*, and *CD79B*. Our data suggest that katagis loci are reasonable indications of aSHM. We observed significantly higher RNA expression of genes with putative aSHM loci compared to those without. In addition, these putative aSHM loci were significantly enriched in genes involved in BCR signaling. Together this implicates that BCR signaling genes are both upregulated and targeted by putative aSHM, raising the question of cause and effect - is aSHM upregulating these genes, or is the high expression levels of these genes priming them for aSHM? This becomes even more complex when considering that highly expressed genes should have lower mutational rates due to transcriptional coupled repair^151^.

The landscape of copy number aberrations and structural variations revealed potentially clinically exploitable deletion of *TOX* as a predictor for anti-PD1 response^107^, amplification of *MALT1*, whose inhibition has been shown to be selectively toxic for ABC-DLBCL^152^, and potential enhancer-hijacking events involving *PIK3C3* and *EPHA4*, whose inhibition has shown therapeutic advantage in a number of cancer models^117-119,124^.

While the genetic landscape of PCNSL was described in some detail before^12-14,16,20,21,39,45,47^, studies investigating the global gene expression profile of PCNSL have been scarce so far. Therefore, we performed RNA sequencing of 37 CNSL samples and 2 normal brain controls. Global gene expression profiles demonstrates that PCNSL are indeed distinct and can be distinguished from systemic ABC-DLBCL. This was perfectly mirrored based on the expression repertoire of *IG* constant genes, implicating the role of B-cell maturation in classification of PCNSL and other lymphomas, as employed in leukemia and multiple myeloma ^153,154^.

PCNSL are highly proliferative^155^. *TERT* activation confers unlimited proliferation, and activating *TERT* promoter mutations are frequent in different types of human cancers^156^. Mutations at two hotspots positions (−124 G>A and −146 G>A) are causal for enhanced *TERT* promoter activity. Bruno *et al*. have previously reported these *TERT* promoter mutations to be present in PCNSL located in the splenium^137^. Therefore, we investigated n = 69 CNSL (including n = 49 PCNSL) tumors, but could not identify any *TERT* promoter mutations suggesting that this mechanisms of *TERT* activation is likely not relevant in PCNSL. Yet, we observed significantly more *TERT* expression in PCNSL compared to non-CNS ABC-DLBCL and this was consistent when stratifying the cohort based on our RNAseq subgrouping. However, we were not able to identify increased telomere content in PCNSL (or MCD) compared to the other groups, suggesting a role of telomere maintenance to overcome telomere shortening, that is imposed by the high levels of proliferation. This concept was supported by a within-group correlation of *TERT* expression and normalised telomere content (Pearson’s R = 0.67, *p* = 0.0023), implicating a role of *TERT* in overcoming telomere degradation in PCNSL. Supporting the high proliferation in these tumors, PCNSL showed significantly elevated presence of the mutational signatures SBS1, which correlates with DNA replication, as well as of ID1 and ID2 that are associated with slippage during DNA replication.

With our study, we have substantiated the genomic and transcriptomic alterations characterizing PCNSL. We show that PCNSL can be clearly distinguished from systemic DLBCL, having distinct expression profiles, *IG* expression and translocation patterns, as well as specific combinations of genetic alterations that are characterized by genomic instability, BCR activation, and most importantly, oncogenic TLR and NFkB signaling, which should be in the focus of future drug development.

## Data Availability

The raw sequencing data of the 51 CNSL samples has been deposited at the European Genome-Phenome Archive under the accession number EGAS00001005339. Accession to the ICGC MMML-Seq data is available via the data accession committee of the ICGC (see: www.ICGC.org, Huebschmann et al). All somatic mutation calls and integrated mutations tables on which the analysis was performed can be downloaded from Zenodo (https://doi.org/10.5281/zenodo.5106351)

## Notes

## Acknowledgements

We thank the German Cancer Research Center (Deutsches Krebsforschungszentrum, DKFZ) Omics IT, Data Management Core Facility (ODCF), and the sequencing unit of the Genomics and Proteomics Core Facility (GPCF) for providing excellent technical support. We thank the DKFZ-Heidelberg Center for Personalized Oncology (DKFZ-HIPO) for technical support and funding through HIPO projects H050 and A050. We thank the German Cancer Consortium (DKTK), Partner site Berlin for technical support and funding through XD013 sequencing. We are indebted to Stefanie Mende, Petra Matylewski, Kathrein Permien, Vera Wolf, Sandra Meier, and Silvia Stefaniak for excellent technical assistance. We thank Prof. Werner Stenzel, PD Dr. Arend Koch, Prof. David Capper, Prof. Christine Sers and Prof. Ingeborg Tinhofer-Keilholz for valuable experimental advice.

## Authors Contribution

JR, RK, ES, FP, SB, DL performed experiments and analysed data. ZG, LS, DH, UT, XPH, NP, MS, MB, SU, NI carried out the bioinformatics analyses and processed data. DP, HR, AO performed sample acquisition and histological analyses. IA performed histological analyses. DL performed FISH analyses. AK, MM, JO, KF, PV, DM, YW, AJ, AU, LT clinically assessed and operated patients and contributed to sample acquisition. CHM, SWi, JR coordinated sample acquisition. SWi, SWo oversaw sequencing of the samples. Experimental design and execution were overseen by FLH, SWi, OW, CHM, BB, CAS, RS, MH. JR, NI, SWi, FLH, RS designed the study. JR and NI wrote the manuscript. JR prepared the final figures. FLH, SWi, OW, CHM, BB, CAS, RS, MH critically revised the manuscript. All authors critically read, reviewed and approved the final manuscript.

## Abbreviations

CNSL: Central nervous system lymphoma
PCNSL: Primary central nervous system lymphoma
SCNSL: Secondary central nervous system lymphoma
ABC: Activated B-cell like
GCB: Germinal center B-cell like
WGS: Whole-genome sequencing
RNAseq: RNA sequencing
H&E: Hematoxylin and eosin
DLBCL: Diffuse large B-cell lymphoma
FL: Follicular lymphoma
MYD88: Myeloid differentiation primary response 88
NFkB: Nuclear factor ‘kappa-light-chain-enhancer’ of activated B-cells
RT-PCR: Real-time PCR
SV direct: Structural variant with breakpoint in the gene body
cnLOH: Copy number neutral loss of heterozygosity
CNA: Copy number aberration
aSHM: Aberrant somatic hyper-mutation
SNV: Single nucleotide variant
indel: Small insertion or deletion
EBV: Epstein-Barr virus
FF: Fresh-frozen
FFPE: Formalin-fixed and paraffin-embedded
VAF: Variant allele frequency
AID: Activation induced deaminase
lncRNA: Long non-coding RNAs

## References

1 Louis, D. N., Ohgaki, H., Wiestler, O. D. & Cavenee, W. K. WHO classification of tumours of the central nervous system. 4 edn, Vol. 1 (International agency for research on cancer Lyon, France, 2016).

2 Swerdlow, S. H. et al. WHO classification of tumours of haematopoietic and lymphoid tissues. Vol. 2 (International agency for research on cancer Lyon, France, 2008).

3 Maciocia, P. et al. Treatment of diffuse large B-cell lymphoma with secondary central nervous system involvement: encouraging efficacy using CNS-penetrating R-IDARAM chemotherapy. British journal of haematology 172, 545–553, doi:10.1111/bjh.13867 (2016).

4 Ferreri, A. J. M. Secondary CNS lymphoma: the poisoned needle in the haystack. Ann Oncol 28, 2335–2337, doi:10.1093/annonc/mdx515 (2017).

5 Montesinos-Rongen, M., Siebert, R. & Deckert, M. Primary lymphoma of the central nervous system: just DLBCL or not? Blood 113, 7–10, doi:10.1182/blood-2008-04-149005 (2009).

6 Grommes, C. & DeAngelis, L. M. Primary CNS Lymphoma. Journal of clinical oncology : official journal of the American Society of Clinical Oncology 35, 2410–2418, doi:10.1200/JCO.2017.72.7602 (2017).

7 Louis, D. N. et al. The 2016 World Health Organization Classification of Tumors of the Central Nervous System: a summary. Acta neuropathologica 131, 803–820, doi:10.1007/s00401-016-1545-1 (2016).

8 Hans, C. P. et al. Confirmation of the molecular classification of diffuse large B-cell lymphoma by immunohistochemistry using a tissue microarray. Blood 103, 275–282, doi:10.1182/blood-2003-05-1545 (2004).

9 Klein, U. et al. Transcriptional analysis of the B cell germinal center reaction. Proceedings of the National Academy of Sciences of the United States of America 100, 2639–2644, doi:10.1073/pnas.0437996100 (2003).

10 Klein, U. et al. Transcription factor IRF4 controls plasma cell differentiation and class-switch recombination. Nature immunology 7, 773–782, doi:10.1038/ni1357 (2006).

11 Montesinos-Rongen, M. et al. Gene expression profiling suggests primary central nervous system lymphomas to be derived from a late germinal center B cell. Leukemia 22, 400–405, doi:10.1038/sj.leu.2405019 (2008).

12 Deckert, M., Montesinos-Rongen, M., Brunn, A. & Siebert, R. Systems biology of primary CNS lymphoma: from genetic aberrations to modeling in mice. Acta neuropathologica 127, 175–188, doi:10.1007/s00401-013-1202-x (2014).

13 Montesinos-Rongen, M. et al. Activating L265P mutations of the MYD88 gene are common in primary central nervous system lymphoma. Acta neuropathologica 122, 791–792, doi:10.1007/s00401-011-0891-2 (2011).

14 Montesinos-Rongen, M. et al. Mutations of CARD11 but not TNFAIP3 may activate the NF-kappaB pathway in primary CNS lymphoma. Acta neuropathologica 120, 529–535, doi:10.1007/s00401-010-0709-7 (2010).

15 Bodor, C. et al. Molecular Subtypes and Genomic Profile of Primary Central Nervous System Lymphoma. Journal of neuropathology and experimental neurology 79, 176–183, doi:10.1093/jnen/nlz125 (2020).

16 Montesinos-Rongen, M., Schafer, E., Siebert, R. & Deckert, M. Genes regulating the B cell receptor pathway are recurrently mutated in primary central nervous system lymphoma. Acta neuropathologica 124, 905–906, doi:10.1007/s00401-012-1064-7 (2012).

17 Jordanova, E. S. et al. Hemizygous deletions in the HLA region account for loss of heterozygosity in the majority of diffuse large B-cell lymphomas of the testis and the central nervous system. Genes Chromosomes Cancer 35, 38–48, doi:10.1002/gcc.10093 (2002).

18 Riemersma, S. A. et al. Extensive genetic alterations of the HLA region, including homozygous deletions of HLA class II genes in B-cell lymphomas arising in immune-privileged sites. Blood 96, 3569–3577 (2000).

19 Montesinos-Rongen, M. et al. Interphase cytogenetic analysis of lymphoma-associated chromosomal breakpoints in primary diffuse large B-cell lymphomas of the central nervous system. Journal of neuropathology and experimental neurology 61, 926–933, doi:10.1093/jnen/61.10.926 (2002).

20 Schwindt, H. et al. Chromosomal translocations fusing the BCL6 gene to different partner loci are recurrent in primary central nervous system lymphoma and may be associated with aberrant somatic hypermutation or defective class switch recombination. Journal of neuropathology and experimental neurology 65, 776–782, doi:10.1097/01.jnen.0000229988.48042.ae (2006).

21 Chapuy, B. et al. Targetable genetic features of primary testicular and primary central nervous system lymphomas. Blood 127, 869–881, doi:10.1182/blood-2015-10-673236 (2016).

22 Gonzalez-Aguilar, A. et al. Recurrent mutations of MYD88 and TBL1XR1 in primary central nervous system lymphomas. Clinical cancer research : an official journal of the American Association for Cancer Research 18, 5203–5211, doi:10.1158/1078-0432.CCR-12-0845 (2012).

23 Kreher, S. et al. Prognostic impact of B-cell lymphoma 6 in primary CNS lymphoma. Neuro-oncology 17, 1016–1021, doi:10.1093/neuonc/nov046 (2015).

24 Riemersma, S. A. et al. High numbers of tumour-infiltrating activated cytotoxic T lymphocytes, and frequent loss of HLA class I and II expression, are features of aggressive B cell lymphomas of the brain and testis. The Journal of pathology 206, 328–336, doi:10.1002/path.1783 (2005).

25 Chapuy, B. et al. Molecular subtypes of diffuse large B cell lymphoma are associated with distinct pathogenic mechanisms and outcomes. Nat Med 24, 679–690, doi:10.1038/s41591-018-0016-8 (2018).

26 Schmitz, R. et al. Genetics and Pathogenesis of Diffuse Large B-Cell Lymphoma. The New England journal of medicine 378, 1396–1407, doi:10.1056/NEJMoa1801445 (2018).

27 Wright, G. W. et al. A Probabilistic Classification Tool for Genetic Subtypes of Diffuse Large B Cell Lymphoma with Therapeutic Implications. Cancer Cell 37, 551–568 e514, doi:10.1016/j.ccell.2020.03.015 (2020).

28 Venturutti, L. & Melnick, A. The dangers of deja vu: Memory B-cells as the cell-of-origin of ABC-DLBCLs. Blood, doi:10.1182/blood.2020005857 (2020).

29 Venturutti, L. et al. TBL1XR1 Mutations Drive Extranodal Lymphoma by Inducing a Protumorigenic Memory Fate. Cell 182, 297–316 e227, doi:10.1016/j.cell.2020.05.049 (2020).

30 Hubschmann, D. et al. Mutational mechanisms shaping the coding and noncoding genome of germinal center derived B-cell lymphomas. Leukemia, doi:10.1038/s41375-021-01251-z (2021).

31 Niparuck, P. et al. Treatment outcome and prognostic factors in PCNSL. Diagn Pathol 14, 56, doi:10.1186/s13000-019-0833-1 (2019).

32 Coiffier, B. et al. CHOP chemotherapy plus rituximab compared with CHOP alone in elderly patients with diffuse large-B-cell lymphoma. The New England journal of medicine 346, 235–242, doi:10.1056/NEJMoa011795 (2002).

33 Schmitt, A. M. et al. Rituximab in primary central nervous system lymphoma-A systematic review and meta-analysis. Hematol Oncol 37, 548–557, doi:10.1002/hon.2666 (2019).

34 Seidel, S. & Schlegel, U. Have treatment protocols for primary CNS lymphoma advanced in the past 10 years. Expert Rev Anticancer Ther 19, 909–915, doi:10.1080/14737140.2019.1677157 (2019).

35 van der Meulen, M. et al. Primary therapy and survival in patients aged over 70-years-old with primary central nervous system lymphoma: a contemporary, nationwide, population-based study in the Netherlands. Haematologica 106, 597–600, doi:10.3324/haematol.2020.247536 (2021).

36 Van Dijck, R., Doorduijn, J. K. & Bromberg, J. E. C. The Role of Rituximab in the Treatment of Primary Central Nervous System Lymphoma. Cancers (Basel) 13, doi:10.3390/cancers13081920 (2021).

37 Deckert, M. et al. Modern concepts in the biology, diagnosis, differential diagnosis and treatment of primary central nervous system lymphoma. Leukemia 25, 1797–1807, doi:10.1038/leu.2011.169 (2011).

38 Braggio, E. et al. Genome-Wide Analysis Uncovers Novel Recurrent Alterations in Primary Central Nervous System Lymphomas. Clinical cancer research : an official journal of the American Association for Cancer Research 21, 3986–3994, doi:10.1158/1078-0432.CCR-14-2116 (2015).

39 Nayyar, N. et al. MYD88 L265P mutation and CDKN2A loss are early mutational events in primary central nervous system diffuse large B-cell lymphomas. Blood Adv 3, 375–383, doi:10.1182/bloodadvances.2018027672 (2019).

40 Grommes, C., Nayak, L., Tun, H. W. & Batchelor, T. T. Introduction of novel agents in the treatment of primary CNS lymphoma. Neuro-oncology 21, 306–313, doi:10.1093/neuonc/noy193 (2019).

41 Ghesquieres, H. et al. Lenalidomide in combination with intravenous rituximab (REVRI) in relapsed/refractory primary CNS lymphoma or primary intraocular lymphoma: a multicenter prospective ‘proof of concept’ phase II study of the French Oculo-Cerebral lymphoma (LOC) Network and the Lymphoma Study Association (LYSA)dagger. Ann Oncol 30, 621–628, doi:10.1093/annonc/mdz032 (2019).

42 Soussain, C. et al. Ibrutinib monotherapy for relapse or refractory primary CNS lymphoma and primary vitreoretinal lymphoma: Final analysis of the phase II ‘proof-of-concept’ iLOC study by the Lymphoma study association (LYSA) and the French oculo-cerebral lymphoma (LOC) network. European journal of cancer 117, 121–130, doi:10.1016/j.ejca.2019.05.024 (2019).

43 Kotla, V. et al. Mechanism of action of lenalidomide in hematological malignancies. J Hematol Oncol 2, 36, doi:10.1186/1756-8722-2-36 (2009).

44 Nayak, L. et al. PD-1 blockade with nivolumab in relapsed/refractory primary central nervous system and testicular lymphoma. Blood 129, 3071–3073, doi:10.1182/blood-2017-01-764209 (2017).

45 Vater, I. et al. The mutational pattern of primary lymphoma of the central nervous system determined by whole-exome sequencing. Leukemia 29, 677–685, doi:10.1038/leu.2014.264 (2015).

46 Bruno, A. et al. Mutational analysis of primary central nervous system lymphoma. Oncotarget 5, 5065–5075, doi:10.18632/oncotarget.2080 (2014).

47 Zhou, Y. et al. Analysis of Genomic Alteration in Primary Central Nervous System Lymphoma and the Expression of Some Related Genes. Neoplasia 20, 1059–1069, doi:10.1016/j.neo.2018.08.012 (2018).

48 Takashima, Y. et al. Target amplicon exome-sequencing identifies promising diagnosis and prognostic markers involved in RTK-RAS and PI3K-AKT signaling as central oncopathways in primary central nervous system lymphoma. Oncotarget 9, 27471–27486, doi:10.18632/oncotarget.25463 (2018).

49 Kaulen, L. D. et al. Whole Exome Sequencing Identifies Novel SLIT2 Mutations in Primary CNS Lymphoma (3962). Neurology 94, 3962 (2020).

50 Gandhi, M. K. et al. EBV-tissue positive primary CNS lymphoma occurring after immunosuppression is a distinct immunobiological entity. Blood, doi:10.1182/blood.2020008520 (2020).

51 Lopez, C. et al. Genomic and transcriptomic changes complement each other in the pathogenesis of sporadic Burkitt lymphoma. Nat Commun 10, 1459, doi:10.1038/s41467-019-08578-3 (2019).

52 Consortium, I. T. P.-C. A. o. W. G. Pan-cancer analysis of whole genomes. Nature 578, 82–93, doi:10.1038/s41586-020-1969-6 (2020).

53 Stocher, M. et al. Parallel detection of five human herpes virus DNAs by a set of real-time polymerase chain reactions in a single run. J Clin Virol 26, 85–93, doi:10.1016/s1386-6532(02)00042-2 (2003).

54 Arber, D. A. et al. The 2016 revision to the World Health Organization classification of myeloid neoplasms and acute leukemia. Blood 127, 2391–2405, doi:10.1182/blood-2016-03-643544 (2016).

55 Leonard, J. P., Martin, P. & Roboz, G. J. Practical Implications of the 2016 Revision of the World Health Organization Classification of Lymphoid and Myeloid Neoplasms and Acute Leukemia. Journal of clinical oncology : official journal of the American Society of Clinical Oncology 35, 2708–2715, doi:10.1200/JCO.2017.72.6745 (2017).

56 Kretzmer, H. et al. DNA methylome analysis in Burkitt and follicular lymphomas identifies differentially methylated regions linked to somatic mutation and transcriptional control. Nature genetics 47, 1316–1325, doi:10.1038/ng.3413 (2015).

57 Li, H. & Durbin, R. Fast and accurate short read alignment with Burrows-Wheeler transform. Bioinformatics 25, 1754–1760, doi:10.1093/bioinformatics/btp324 (2009).

58 Tischler, G. & Leonard, S. biobambam: tools for read pair collation based algorithms on BAM files. Source Code for Biology and Medicine 9, 13, doi:10.1186/1751-0473-9-13 (2014).

59 Ishaque, N. et al. Whole genome sequencing puts forward hypotheses on metastasis evolution and therapy in colorectal cancer. Nat Commun 9, 4782, doi:10.1038/s41467-018-07041-z (2018).

60 Li, H. et al. The Sequence Alignment/Map format and SAMtools. Bioinformatics 25, 2078–2079, doi:10.1093/bioinformatics/btp352 (2009).

61 Consortium, E. P. An integrated encyclopedia of DNA elements in the human genome. Nature 489, 57–74, doi:10.1038/nature11247 (2012).

62 Rimmer, A. et al. Integrating mapping-, assembly- and haplotype-based approaches for calling variants in clinical sequencing applications. Nature genetics 46, 912–918, doi:10.1038/ng.3036 (2014).

63 Wang, K., Li, M. & Hakonarson, H. ANNOVAR: functional annotation of genetic variants from high-throughput sequencing data. Nucleic acids research 38, e164, doi:10.1093/nar/gkq603 (2010).

64 Jiang, Y., Qiu, Y., Minn, A. J. & Zhang, N. R. Assessing intratumor heterogeneity and tracking longitudinal and spatial clonal evolutionary history by next-generation sequencing. Proceedings of the National Academy of Sciences of the United States of America 113, E5528–5537, doi:10.1073/pnas.1522203113 (2016).

65 Sahm, F. et al. Meningiomas induced by low-dose radiation carry structural variants of NF2 and a distinct mutational signature. Acta neuropathologica 134, 155–158, doi:10.1007/s00401-017-1715-9 (2017).

66 Uhrig, S. et al. Accurate and efficient detection of gene fusions from RNA sequencing data. Genome Res 31, 448–460, doi:10.1101/gr.257246.119 (2021).

67 Kleinheinz, K. et al. ACEseq – allele specific copy number estimation from whole genome sequencing. bioRxiv, 210807, doi:10.1101/210807 (2017).

68 Alexandrov, L. B. et al. Signatures of mutational processes in human cancer. Nature 500, 415–421, doi:10.1038/nature12477 (2013).

69 Hubschmann, D. et al. Analysis of mutational signatures with yet another package for signature analysis. Genes Chromosomes Cancer, doi:10.1002/gcc.22918 (2020).

70 Alexandrov, L. B. et al. The repertoire of mutational signatures in human cancer. Nature 578, 94–101, doi:10.1038/s41586-020-1943-3 (2020).

71 Leiserson, M. D., Wu, H. T., Vandin, F. & Raphael, B. J. CoMEt: a statistical approach to identify combinations of mutually exclusive alterations in cancer. Genome biology 16, 160, doi:10.1186/s13059-015-0700-7 (2015).

72 Gonzalez-Perez, A. et al. IntOGen-mutations identifies cancer drivers across tumor types. Nature methods 10, 1081–1082, doi:10.1038/nmeth.2642 (2013).

73 Feuerbach, L. et al. TelomereHunter - in silico estimation of telomere content and composition from cancer genomes. BMC Bioinformatics 20, 272, doi:10.1186/s12859-019-2851-0 (2019).

74 Paramasivam, N. et al. Mutational patterns and regulatory networks in epigenetic subgroups of meningioma. Acta neuropathologica 138, 295–308, doi:10.1007/s00401-019-02008-w (2019).

75 Dobin, A. et al. STAR: ultrafast universal RNA-seq aligner. Bioinformatics 29, 15–21, doi:10.1093/bioinformatics/bts635 (2013).

76 Liao, Y., Smyth, G. K. & Shi, W. featureCounts: an efficient general purpose program for assigning sequence reads to genomic features. Bioinformatics 30, 923–930, doi:10.1093/bioinformatics/btt656 (2014).

77 Gu, Z., Schlesner, M. & Hubschmann, D. cola: an R/Bioconductor package for consensus partitioning through a general framework. Nucleic acids research 49, e15, doi:10.1093/nar/gkaa1146 (2021).

78 Radke, J. et al. Predictive MGMT status in a homogeneous cohort of IDH wildtype glioblastoma patients. Acta neuropathologica communications 7, 89, doi:10.1186/s40478-019-0745-z (2019).

79 Jurmeister, P. et al. Parallel screening for ALK, MET and ROS1 alterations in non-small cell lung cancer with implications for daily routine testing. Lung Cancer 87, 122–129, doi:10.1016/j.lungcan.2014.11.018 (2015).

80 Zhou, Y. et al. Metascape provides a biologist-oriented resource for the analysis of systems-level datasets. Nat Commun 10, 1523, doi:10.1038/s41467-019-09234-6 (2019).

81 Yang, X. et al. STAT3 Activation Is Associated with Interleukin-10 Expression and Survival in Primary Central Nervous System Lymphoma. World Neurosurg 134, e1077–e1084, doi:10.1016/j.wneu.2019.11.100 (2020).

82 Tang, D. et al. Clinicopathologic significance of MYD88 L265P mutation and expression of TLR4 and P-STAT3 in primary central nervous system diffuse large B-cell lymphomas. Brain Tumor Pathol 38, 50–58, doi:10.1007/s10014-020-00386-8 (2021).

83 Bai, L. et al. A Potent and Selective Small-Molecule Degrader of STAT3 Achieves Complete Tumor Regression In Vivo. Cancer Cell 36, 498–511 e417, doi:10.1016/j.ccell.2019.10.002 (2019).

84 Komohara, Y. et al. M2 macrophage/microglial cells induce activation of Stat3 in primary central nervous system lymphoma. J Clin Exp Hematop 51, 93–99, doi:10.3960/jslrt.51.93 (2011).

85 Ngo, V. N. et al. Oncogenically active MYD88 mutations in human lymphoma. Nature 470, 115–119, doi:10.1038/nature09671 (2011).

86 Wang, J. Q. et al. Synergistic cooperation and crosstalk between MYD88(L265P) and mutations that dysregulate CD79B and surface IgM. The Journal of experimental medicine 214, 2759–2776, doi:10.1084/jem.20161454 (2017).

87 Visco, C. et al. Oncogenic Mutations of MYD88 and CD79B in Diffuse Large B-Cell Lymphoma and Implications for Clinical Practice. Cancers (Basel) 12, doi:10.3390/cancers12102913 (2020).

88 Reddy, A. et al. Genetic and Functional Drivers of Diffuse Large B Cell Lymphoma. Cell 171, 481–494 e415, doi:10.1016/j.cell.2017.09.027 (2017).

89 Amodio, N. et al. MALAT1: a druggable long non-coding RNA for targeted anti-cancer approaches. J Hematol Oncol 11, 63, doi:10.1186/s13045-018-0606-4 (2018).

90 Li, S., Li, J., Chen, C., Zhang, R. & Wang, K. Pan-cancer analysis of long non-coding RNA NEAT1 in various cancers. Genes Dis 5, 27–35, doi:10.1016/j.gendis.2017.11.003 (2018).

91 Fujimoto, A. et al. Whole-genome mutational landscape and characterization of noncoding and structural mutations in liver cancer. Nature genetics 48, 500–509, doi:10.1038/ng.3547 (2016).

92 Zheng, Z. H., You, H. Y., Feng, Y. J. & Zhang, Z. T. LncRNA KCNQ1OT1 is a key factor in the reversal effect of curcumin on cisplatin resistance in the colorectal cancer cells. Mol Cell Biochem, doi:10.1007/s11010-020-03856-x (2020).

93 Xu, B. et al. LncRNA SNHG3, a potential oncogene in human cancers. Cancer Cell Int 20, 536, doi:10.1186/s12935-020-01608-x (2020).

94 Tian, Y. et al. lncRNA SNHG14 promotes oncogenesis and immune evasion in diffuse large-B-cell lymphoma by sequestering miR-152-3p. Leuk Lymphoma, 1–15, doi:10.1080/10428194.2021.1876866 (2021).

95 Casellas, R. et al. Mutations, kataegis and translocations in B cells: understanding AID promiscuous activity. Nature reviews. Immunology 16, 164–176, doi:10.1038/nri.2016.2 (2016).

96 Montesinos-Rongen, M., Van Roost, D., Schaller, C., Wiestler, O. D. & Deckert, M. Primary diffuse large B-cell lymphomas of the central nervous system are targeted by aberrant somatic hypermutation. Blood 103, 1869–1875, doi:10.1182/blood-2003-05-1465 (2004).

97 Khodabakhshi, A. H. et al. Recurrent targets of aberrant somatic hypermutation in lymphoma. Oncotarget 3, 1308–1319, doi:10.18632/oncotarget.653 (2012).

98 Papavasiliou, F. N. & Schatz, D. G. Somatic hypermutation of immunoglobulin genes: merging mechanisms for genetic diversity. Cell 109 Suppl, S35–44, doi:10.1016/s0092-8674(02)00706-7 (2002).

99 Morin, R. D. et al. Mutational and structural analysis of diffuse large B-cell lymphoma using whole-genome sequencing. Blood 122, 1256–1265, doi:10.1182/blood-2013-02-483727 (2013).

100 Rimsza, L. M. et al. Loss of MHC class II gene and protein expression in diffuse large B-cell lymphoma is related to decreased tumor immunosurveillance and poor patient survival regardless of other prognostic factors: a follow-up study from the Leukemia and Lymphoma Molecular Profiling Project. Blood 103, 4251–4258, doi:10.1182/blood-2003-07-2365 (2004).

101 Baruah, P. et al. Impact of p16 status on pro- and anti-angiogenesis factors in head and neck cancers. British journal of cancer 113, 653–659, doi:10.1038/bjc.2015.251 (2015).

102 Eymin, B., Leduc, C., Coll, J. L., Brambilla, E. & Gazzeri, S. p14ARF induces G2 arrest and apoptosis independently of p53 leading to regression of tumours established in nude mice. Oncogene 22, 1822–1835, doi:10.1038/sj.onc.1206303 (2003).

103 England, N. L. et al. Identification of human tumour suppressor genes by monochromosome transfer: rapid growth-arrest response mapped to 9p21 is mediated solely by the cyclin-D-dependent kinase inhibitor gene, CDKN2A (p16INK4A). Carcinogenesis 17, 1567–1575, doi:10.1093/carcin/17.8.1567 (1996).

104 Challa-Malladi, M. et al. Combined genetic inactivation of beta2-Microglobulin and CD58 reveals frequent escape from immune recognition in diffuse large B cell lymphoma. Cancer Cell 20, 728–740, doi:10.1016/j.ccr.2011.11.006 (2011).

105 Schwindt, H. et al. Chromosomal imbalances and partial uniparental disomies in primary central nervous system lymphoma. Leukemia 23, 1875–1884, doi:10.1038/leu.2009.120 (2009).

106 Yu, X. & Li, Z. TOX gene: a novel target for human cancer gene therapy. Am J Cancer Res 5, 3516–3524 (2015).

107 Kim, K. et al. Single-cell transcriptome analysis reveals TOX as a promoting factor for T-cell exhaustion and a predictor for anti-PD1 responses in human cancer. bioRxiv, 641316, doi:10.1101/641316 (2019).

108 Astuti, D. et al. Germline mutations in DIS3L2 cause the Perlman syndrome of overgrowth and Wilms tumor susceptibility. Nature genetics 44, 277–284, doi:10.1038/ng.1071 (2012).

109 Xing, S. et al. DIS3L2 Promotes Progression of Hepatocellular Carcinoma via hnRNP U-Mediated Alternative Splicing. Cancer Res 79, 4923–4936, doi:10.1158/0008-5472.CAN-19-0376 (2019).

110 McAllister-Lucas, L. M., Baens, M. & Lucas, P. C. MALT1 protease: a new therapeutic target in B lymphoma and beyond? Clinical cancer research : an official journal of the American Association for Cancer Research 17, 6623–6631, doi:10.1158/1078-0432.CCR-11-0467 (2011).

111 Kameoka, Y. et al. Contig array CGH at 3p14.2 points to the FRA3B/FHIT common fragile region as the target gene in diffuse large B-cell lymphoma. Oncogene 23, 9148–9154, doi:10.1038/sj.onc.1208136 (2004).

112 Roy, D., Sin, S. H., Damania, B. & Dittmer, D. P. Tumor suppressor genes FHIT and WWOX are deleted in primary effusion lymphoma (PEL) cell lines. Blood 118, e32–39, doi:10.1182/blood-2010-12-323659 (2011).

113 Haller, F. et al. Enhancer hijacking activates oncogenic transcription factor NR4A3 in acinic cell carcinomas of the salivary glands. Nat Commun 10, 368, doi:10.1038/s41467-018-08069-x (2019).

114 Zhang, Y. et al. High-coverage whole-genome analysis of 1220 cancers reveals hundreds of genes deregulated by rearrangement-mediated cis-regulatory alterations. Nat Commun 11, 736, doi:10.1038/s41467-019-13885-w (2020).

115 Zhou, X., Takatoh, J. & Wang, F. The mammalian class 3 PI3K (PIK3C3) is required for early embryogenesis and cell proliferation. PloS one 6, e16358, doi:10.1371/journal.pone.0016358 (2011).

116 Munson, M. J. & Ganley, I. G. MTOR, PIK3C3, and autophagy: Signaling the beginning from the end. Autophagy 11, 2375–2376, doi:10.1080/15548627.2015.1106668 (2015).

117 Chen, C. H. et al. Dual Inhibition of PIK3C3 and FGFR as a New Therapeutic Approach to Treat Bladder Cancer. Clinical cancer research : an official journal of the American Association for Cancer Research 24, 1176–1189, doi:10.1158/1078-0432.CCR-17-2066 (2018).

118 Kumar, B. et al. PIK3C3 Inhibition Promotes Sensitivity to Colon Cancer Therapy by Inhibiting Cancer Stem Cells. Cancers (Basel) 13, doi:10.3390/cancers13092168 (2021).

119 Liu, F. et al. PIK3C3 regulates the expansion of liver CSCs and PIK3C3 inhibition counteracts liver cancer stem cell activity induced by PI3K inhibitor. Cell Death Dis 11, 427, doi:10.1038/s41419-020-2631-9 (2020).

120 Fukai, J. et al. EphA4 promotes cell proliferation and migration through a novel EphA4-FGFR1 signaling pathway in the human glioma U251 cell line. Mol Cancer Ther 7, 2768–2778, doi:10.1158/1535-7163.MCT-07-2263 (2008).

121 Iiizumi, M. et al. EphA4 receptor, overexpressed in pancreatic ductal adenocarcinoma, promotes cancer cell growth. Cancer Sci 97, 1211–1216, doi:10.1111/j.1349-7006.2006.00313.x (2006).

122 Hachim, I. Y. et al. Transforming Growth Factor-beta Regulation of Ephrin Type-A Receptor 4 Signaling in Breast Cancer Cellular Migration. Scientific reports 7, 14976, doi:10.1038/s41598-017-14549-9 (2017).

123 Lin, C. Y. et al. High Expression of EphA4 Predicted Lesser Degree of Tumor Regression after Neoadjuvant Chemoradiotherapy in Rectal Cancer. J Cancer 8, 1089–1096, doi:10.7150/jca.17471 (2017).

124 Kina, S. et al. Targeting EphA4 abrogates intrinsic resistance to chemotherapy in well-differentiated cervical cancer cell line. Eur J Pharmacol 840, 70–78, doi:10.1016/j.ejphar.2018.09.031 (2018).

125 Schmitz, R., Ceribelli, M., Pittaluga, S., Wright, G. & Staudt, L. M. Oncogenic mechanisms in Burkitt lymphoma. Cold Spring Harb Perspect Med 4, doi:10.1101/cshperspect.a014282 (2014).

126 Kuppers, R. & Dalla-Favera, R. Mechanisms of chromosomal translocations in B cell lymphomas. Oncogene 20, 5580–5594, doi:10.1038/sj.onc.1204640 (2001).

127 Seifert, M., Scholtysik, R. & Kuppers, R. Origin and Pathogenesis of B Cell Lymphomas. Methods in molecular biology 1956, 1–33, doi:10.1007/978-1-4939-9151-8_1 (2019).

128 Krull, J. E. et al. Somatic copy number gains in MYC, BCL2, and BCL6 identifies a subset of aggressive alternative-DH/TH DLBCL patients. Blood Cancer J 10, 117, doi:10.1038/s41408-020-00382-3 (2020).

129 Scott, D. W. et al. High-grade B-cell lymphoma with MYC and BCL2 and/or BCL6 rearrangements with diffuse large B-cell lymphoma morphology. Blood 131, 2060–2064, doi:10.1182/blood-2017-12-820605 (2018).

130 Fong, P. C. et al. Inhibition of poly(ADP-ribose) polymerase in tumors from BRCA mutation carriers. The New England journal of medicine 361, 123–134, doi:10.1056/NEJMoa0900212 (2009).

131 Poti, A. et al. Correlation of homologous recombination deficiency induced mutational signatures with sensitivity to PARP inhibitors and cytotoxic agents. Genome biology 20, 240, doi:10.1186/s13059-019-1867-0 (2019).

132 Seimiya, M. et al. Stage-specific expression of Clast6/E3/LAPTM5 during B cell differentiation: elevated expression in human B lymphomas. Int J Oncol 22, 301–304 (2003).

133 Kim, Y., Shin, Y. & Kang, G. H. Prognostic significance of CD103+ immune cells in solid tumor: a systemic review and meta-analysis. Scientific reports 9, 3808, doi:10.1038/s41598-019-40527-4 (2019).

134 Harada, K. et al. Telomerase activity in central nervous system malignant lymphoma. Cancer 86, 1050–1055 (1999).

135 Panebianco, F., Nikitski, A. V., Nikiforova, M. N. & Nikiforov, Y. E. Spectrum of TERT promoter mutations and mechanisms of activation in thyroid cancer. Cancer Med 8, 5831–5839, doi:10.1002/cam4.2467 (2019).

136 Bell, R. J. et al. Understanding TERT Promoter Mutations: A Common Path to Immortality. Mol Cancer Res 14, 315–323, doi:10.1158/1541-7786.MCR-16-0003 (2016).

137 Bruno, A. et al. TERT promoter mutations in primary central nervous system lymphoma are associated with spatial distribution in the splenium. Acta neuropathologica 130, 439–440, doi:10.1007/s00401-015-1461-9 (2015).

138 Stogbauer, L., Stummer, W., Senner, V. & Brokinkel, B. Telomerase activity, TERT expression, hTERT promoter alterations, and alternative lengthening of the telomeres (ALT) in meningiomas - a systematic review. Neurosurg Rev, doi:10.1007/s10143-019-01087-3 (2019).

139 Ichimura, K. TERT promoter mutation as a diagnostic marker for diffuse gliomas. Neurooncology 21, 417–418, doi:10.1093/neuonc/noz025 (2019).

140 Lee, Y. et al. The frequency and prognostic effect of TERT promoter mutation in diffuse gliomas. Acta neuropathologica communications 5, 62, doi:10.1186/s40478-017-0465-1 (2017).

141 Liu, Z. et al. Association between TERT rs2853669 polymorphism and cancer risk: A meta-analysis of 9,157 cases and 11,073 controls. PloS one 13, e0191560, doi:10.1371/journal.pone.0191560 (2018).

142 Grommes, C. & Younes, A. Ibrutinib in PCNSL: The Curious Cases of Clinical Responses and Aspergillosis. Cancer Cell 31, 731–733, doi:10.1016/j.ccell.2017.05.004 (2017).

143 Rheinbay, E. et al. Analyses of non-coding somatic drivers in 2,658 cancer whole genomes. Nature 578, 102–111, doi:10.1038/s41586-020-1965-x (2020).

144 Biesecker, L. G., Shianna, K. V. & Mullikin, J. C. Exome sequencing: the expert view. Genome biology 12, 128, doi:10.1186/gb-2011-12-9-128 (2011).

145 Fatica, A. & Bozzoni, I. Long non-coding RNAs: new players in cell differentiation and development. Nat Rev Genet 15, 7–21, doi:10.1038/nrg3606 (2014).

146 Jardin, F. et al. Diffuse large B-cell lymphomas with CDKN2A deletion have a distinct gene expression signature and a poor prognosis under R-CHOP treatment: a GELA study. Blood 116, 1092–1104, doi:10.1182/blood-2009-10-247122 (2010).

147 Fangazio, M. et al. Genetic mechanisms of HLA-I loss and immune escape in diffuse large B cell lymphoma. Proceedings of the National Academy of Sciences of the United States of America 118, doi:10.1073/pnas.2104504118 (2021).

148 Monti, S. et al. Integrative analysis reveals an outcome-associated and targetable pattern of p53 and cell cycle deregulation in diffuse large B cell lymphoma. Cancer Cell 22, 359–372, doi:10.1016/j.ccr.2012.07.014 (2012).

149 Tao, W. & Levine, A. J. P19(ARF) stabilizes p53 by blocking nucleo-cytoplasmic shuttling of Mdm2. Proceedings of the National Academy of Sciences of the United States of America 96, 6937–6941, doi:10.1073/pnas.96.12.6937 (1999).

150 Yamada, S., Ishida, Y., Matsuno, A. & Yamazaki, K. Primary diffuse large B-cell lymphomas of central nervous system exhibit remarkably high prevalence of oncogenic MYD88 and CD79B mutations. Leuk Lymphoma 56, 2141–2145, doi:10.3109/10428194.2014.979413 (2015).

151 Georgakopoulos-Soares, I. et al. Transcription-coupled repair and mismatch repair contribute towards preserving genome integrity at mononucleotide repeat tracts. Nat Commun 11, 1980, doi:10.1038/s41467-020-15901-w (2020).

152 Ferch, U. et al. Inhibition of MALT1 protease activity is selectively toxic for activated B cell-like diffuse large B cell lymphoma cells. The Journal of experimental medicine 206, 2313–2320, doi:10.1084/jem.20091167 (2009).

153 Oakes, C. C. et al. DNA methylation dynamics during B cell maturation underlie a continuum of disease phenotypes in chronic lymphocytic leukemia. Nature genetics 48, 253–264, doi:10.1038/ng.3488 (2016).

154 Bodker, J. S. et al. A multiple myeloma classification system that associates normal B-cell subset phenotypes with prognosis. Blood Adv 2, 2400–2411, doi:10.1182/bloodadvances.2018018564 (2018).

155 Sugita, Y. et al. Primary central nervous system lymphomas and related diseases: Pathological characteristics and discussion of the differential diagnosis. Neuropathology 36, 313–324, doi:10.1111/neup.12276 (2016).

156 Vinagre, J. et al. Frequency of TERT promoter mutations in human cancers. Nat Commun 4, 2185, doi:10.1038/ncomms3185 (2013).

