## Supplementary figures and images for "The genomic and transcriptional landscape of primary central nervous system lymphoma"

### Supplementary Figures

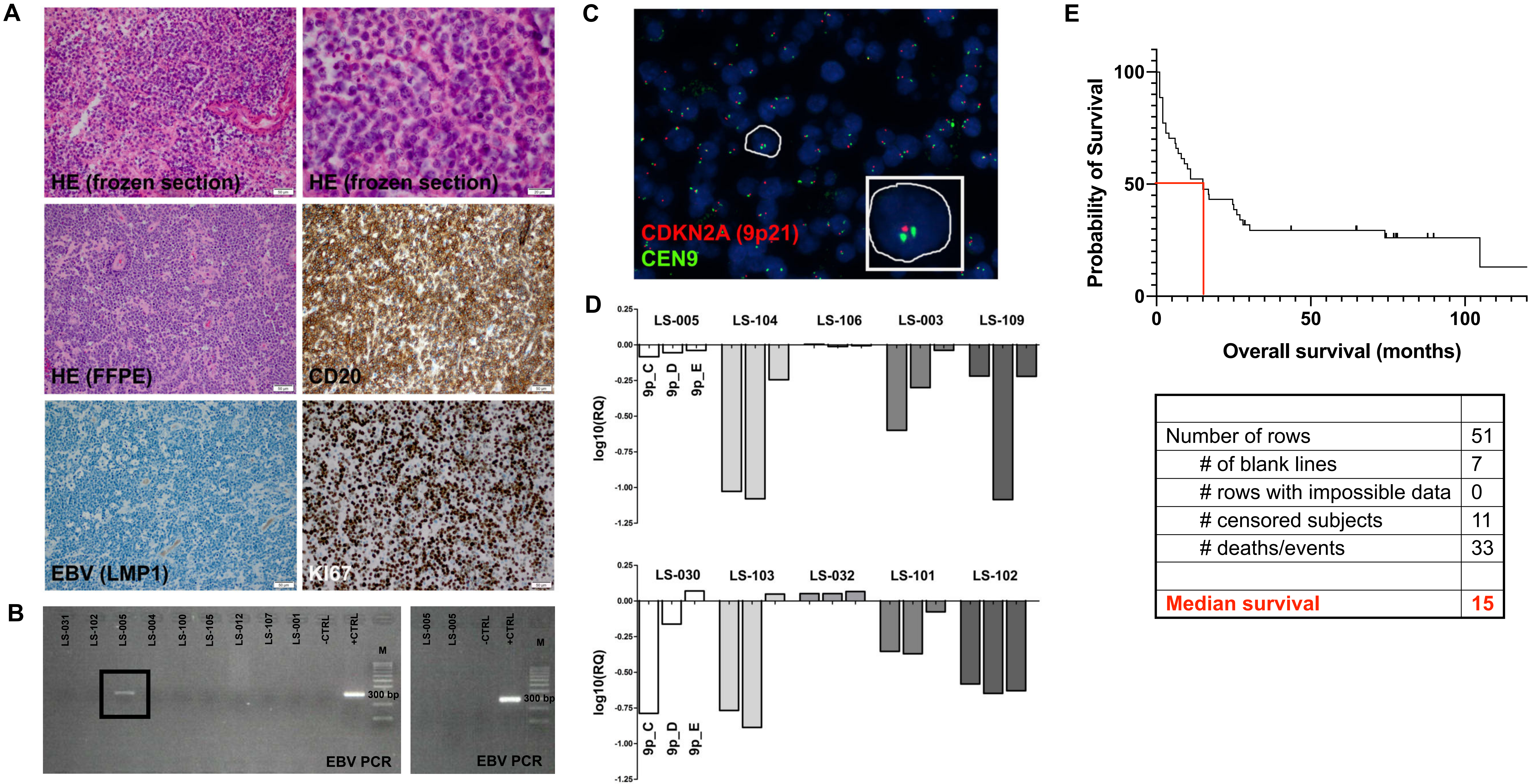

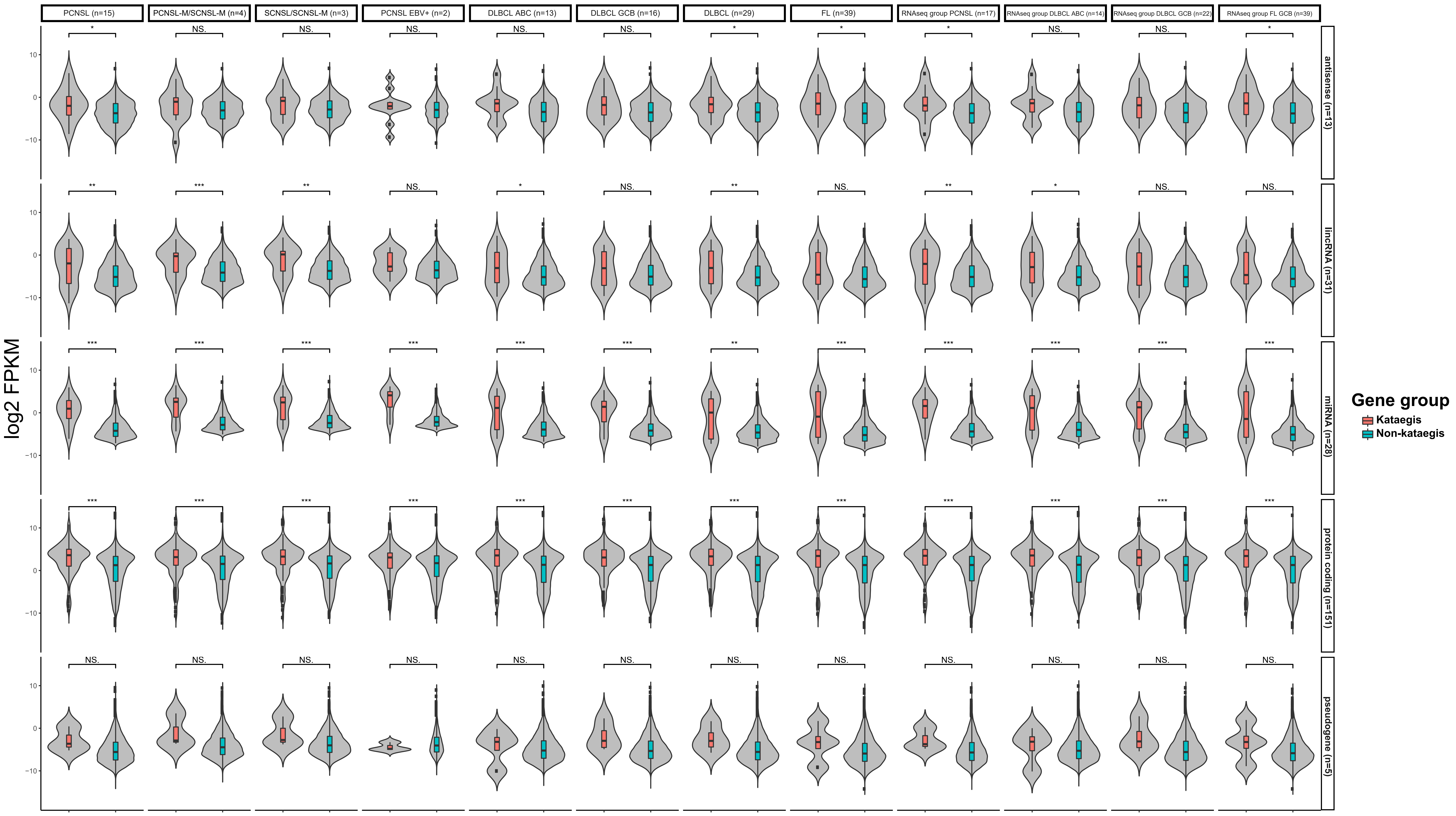

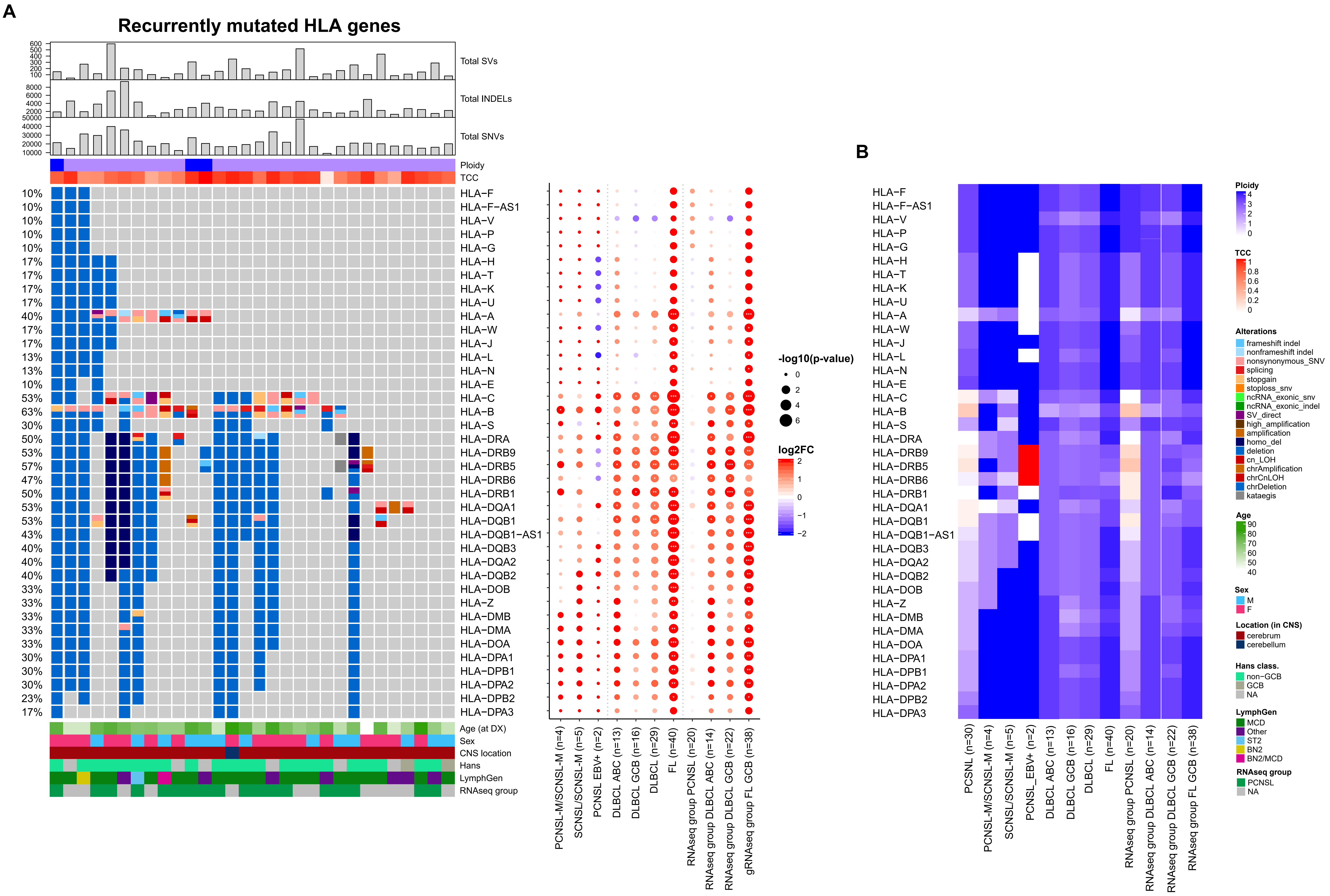

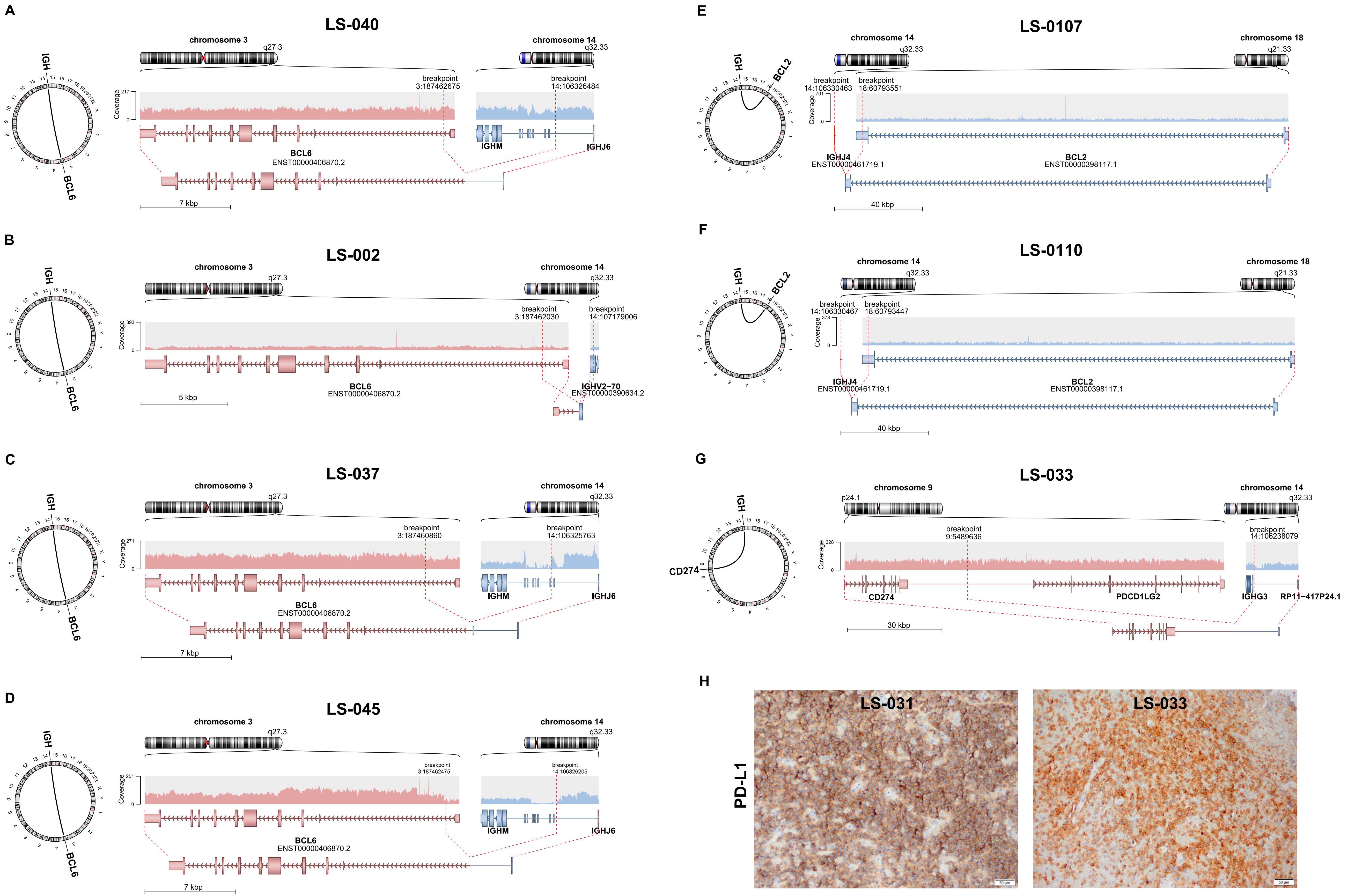

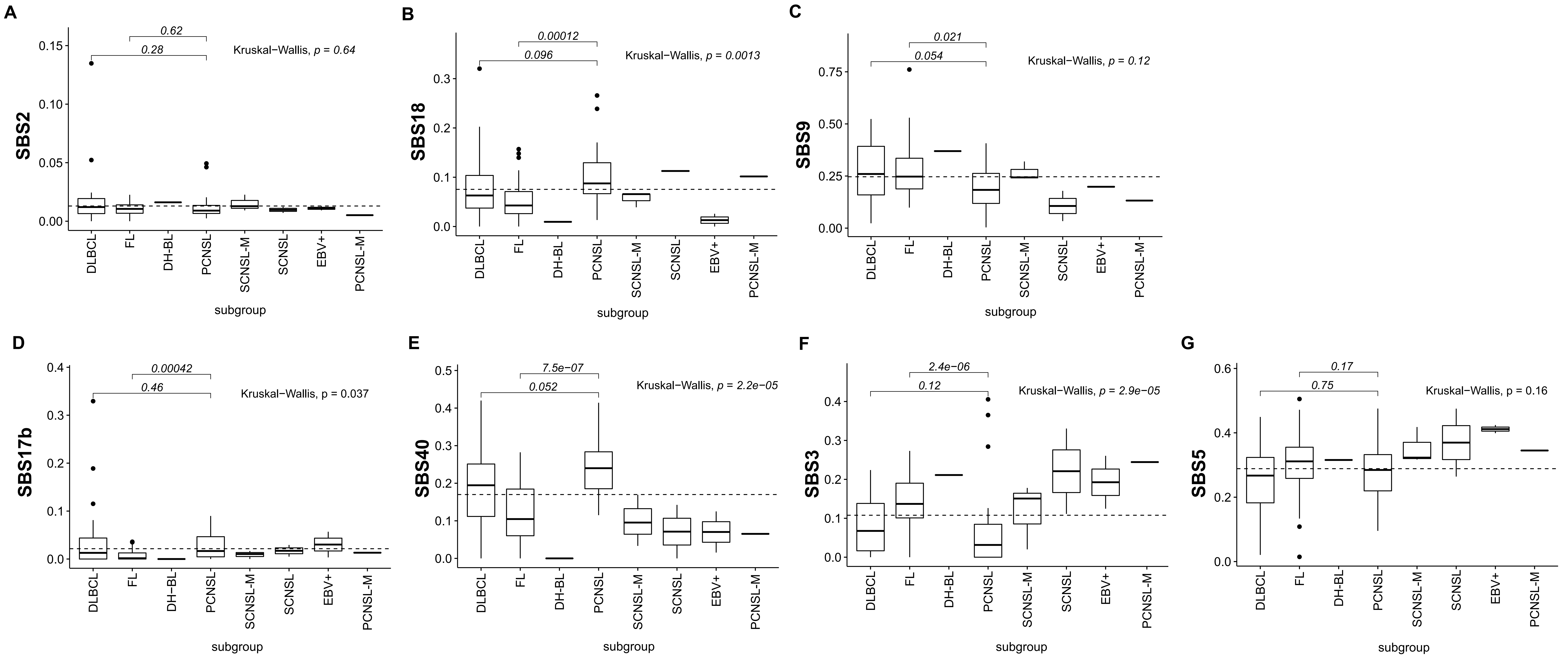

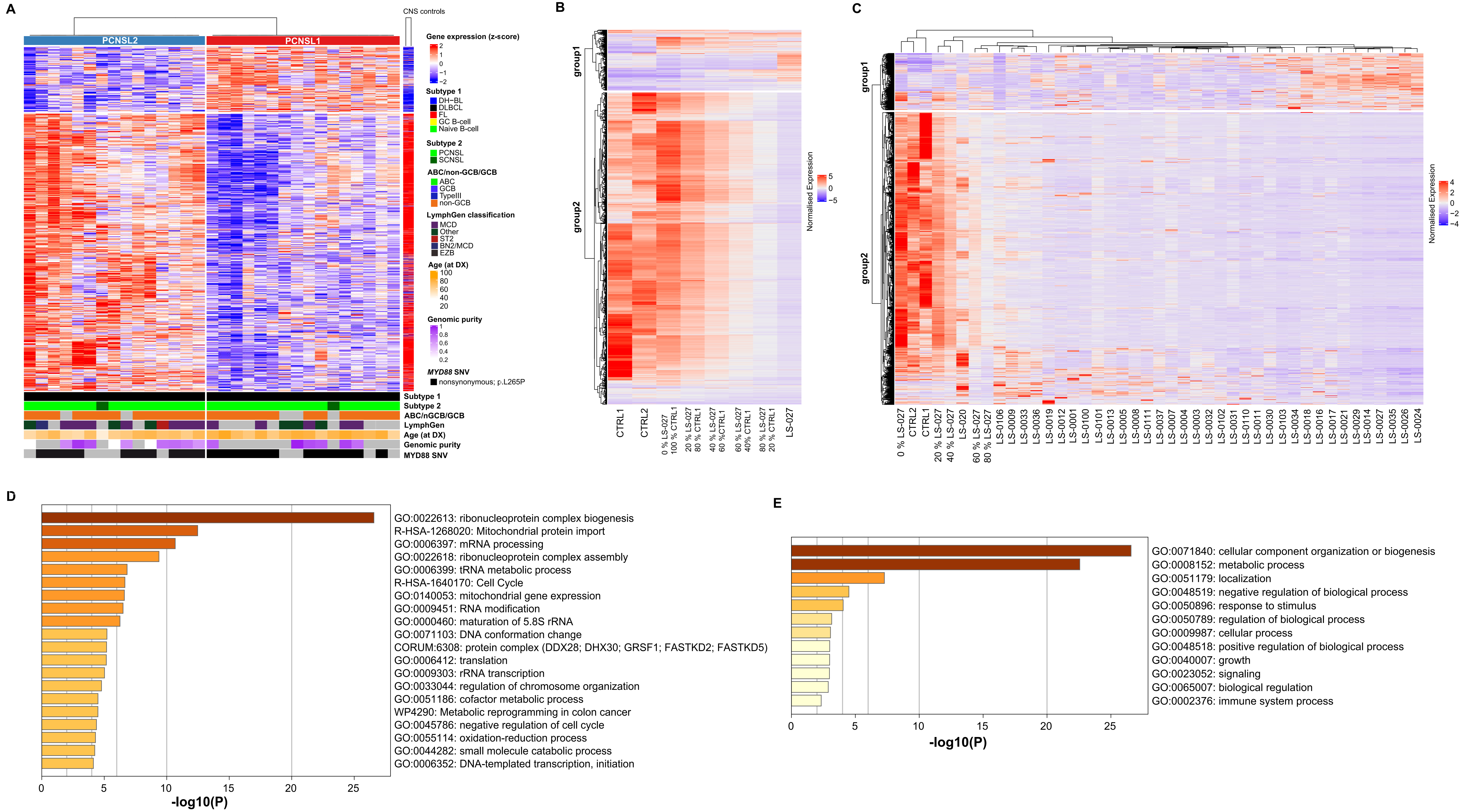

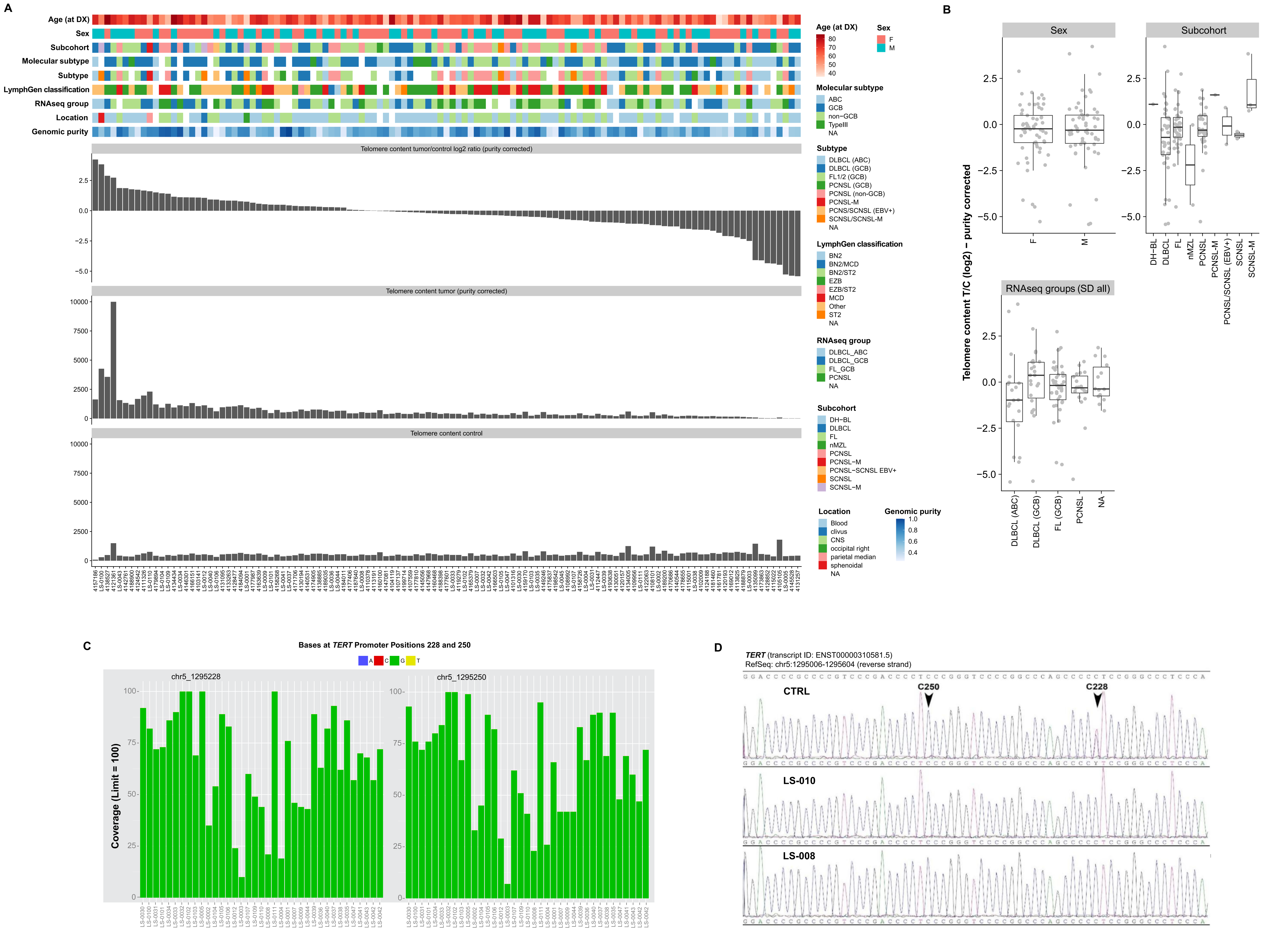
