## Supplementary Figure Legends for "The genomic and transcriptional landscape of primary central nervous system lymphoma"

Supplementary figure 1: **(A)** Histological and immunohistological evaluation of primary CNS lymphoma samples. Hematoxylin & Eosin (H&E) was used to stain frozen sections for histological analysis of intraoperative tissue specimens to evaluate tumour cell content and tissue quality. Formalin-fixed paraffin-embedded (FFPE) samples showed a dense infiltrate of malignant, abnormal large lymphoid cells with vesicular chromatin, prominent nucleoli and cuffing of the capillary vessels. The diagnosis was confirmed by positive immunohistochemistry for CD20. The latent membrane protein 1 (LMP1) of Epstein-Barr virus was not expressed and Ki67 showed very high proliferative activity (> 80%) of the lymphoma cells. **(B)** Detection of Epstein-Barr virus (EBV) DNA in CNSL specimens by PCR targeting a highly conserved region of the EBNA-1 (BKRF1) gene (297 bp, upper panel). Sample LS-GD-005 was positive in a first PCR run (left, black square) but negative in a repeated second run (right). M (1 kb DNA ladder), +CTRL (positive control), -CTRL (negative control; water). **(C)** Exemplary result of CNV validation experiments by fluorescence in situ hybridization (FISH). FISH analysis with a p16 (*CDKN2A*) and CEN9 probe revealed heterozygous deletion with loss of one of the red (*CDKN2A*) signals in a nucleus and preservation of the two centromeric green (CEN9) signals (patient LS-031). **(D)** Quantitative RT-PCR demonstrated homozygous deletion of *CDKN2A* in patients LS-104, LS-003, LS-109, LS-030, LS-103, LS-101, and LS-102 and no *CDKN2A* deletion in patients LS-005, LS-106, and LS-032 using three different primer (9p21.3_C, _D, _E) for the region of *CDKN2A*. **(E)** Kaplan-Meier survival curve analysis (GraphPad Prism 9, version 9.0.0) for the CNSL cohort. Follow up data was available for 44 patients. The follow up time ranged from one to 104 months with a median survival of 15.0 months (red line). Censored subjects are indicated on the Kaplan-Meier curve as tick marks.

Supplementary figure 2: The violin plot shows the RNA expression of genes with kataegis loci compared to those without for antisense, long non-coding RNA, miRNA and protein coding genes for all subcohorts and RNA subgroups (Wilcoxon rank sum test, $p < 0.05$).

Supplementary figure 3: **(A)** Oncoprint of recurrently mutated HLA genes in PCNSL. The top panel of the oncoprint shows the total numbers of structural variants (SVs), small insertions/deletions (INDELs), single nucleotide variants (SNVs), estimated ploidy, and tumour purity. Mutated genes are listed from top to bottom depending on their chromosome location. The colour of the box indicates the type of mutation. The corresponding dot plot reflects the log2 fold change and significance of alteration frequencies in the other subcohorts and RNAseq subgroups compared to PCNSL. The size of the dots demonstrate the significance according to a 2-tailed Fisher´s exact test. **(B)** The heatmaps reflect the alteration frequency of each gene in the other subcohort, RNAseq groups, and LymphGen groups.

Supplementary figure 4: Schematic representation of the translocation breakpoints involving *BCL6* (PCNSL patients LS-040 **(A)**, LS-002 **(B)**, LS-037 **(C)**, LS-045 **(D)**), *BCL2* (SCNSL patient LS-0107 **(E)**, PCNSL-M patient LS-0110 **(F)**), and *CD274* (PD-L1; PCNSL patient LS-033, **(G)**). Immunohistochmical staining revealed PD-L1 expression in both PCNSL patients with translocations involving *CD274* (LS-031, LS-033 **(H)**).

Supplementary figure 5: Statistical analysis of SBS signatures in CNSL and peripheral lymphoma. Shown are the results of nonparametric Kruskal-Wallis H test and pairwise comparison of PCNSL with DLBCL or FL (Mann-Whitney U test) for signatures SBS2 **(A)**, SBS18 **(B)**, SBS9 **(C)**, SBS17b **(D)**, SBS40 **(E)**, SBS3 **(F)**, and SBS5 **(G).**

Supplementary figure 6: **(A)** RNA sequencing was performed using normal brain tissue (frontal lobe) controls (CTRL 1, 2) to extract the brain tissue signature from the PCNSL signature and to investigate the impact of normal brain tissue contamination in PCNSL samples. The heatmap shows unsupervised consensus clustering (using cola with "ATC" as top-value method), which revealed two groups: PCNSL1, (“pure”, right) consisted of samples with high tumor cell content, and PCNSL2 (“impure”, left) contained mainly samples with a lower tumor cell content, which signatures correlated well with normal brain tissue expression. **(B)** We mixed a pure PCNSL sample (LS-027) with increasing concentrations of RNA isolated from normal CNS tissue (CTRL 1, ranging from 0% to 80%). **(C)** The heatmap illustrate the impact of CNS tissue contamination in total RNA sequencing analysis of the tumour tissue. The differentially expressed genes of the pure PCNSL groups were analysed by Metascape[1] to identify the enriched pathways **(D)** and top three level Gene Ontology biological processes **(E)**.

Supplementary figure 7: **(A)** Telomere content in DLBCL, FL, PCNSL and SCNSL was estimated with TelomereHunter using default settings (filtering of telomere reads: at least six telomere repeats per 100 bp read length). In about 1/3 of the samples, the telomere content was higher in the tumor than in the control sample. For samples with information on tumor purity, the telomere content of the tumor sample was corrected. Therefore, the telomere content of the blood control was used as an approximation for that of normal cells. **(B)** There was no statistical difference between PCNSL and systemic DLBCL in telomere content. Comparison of telomere content of different subgroups considering e.g. sex, subcohort, and RNAseq groups did not reach significance. **(C)** We detected no *TERT* promotor mutation in the whole-genome sequencing cohort. The coverage was relatively high in most cases. If there was a mutation, the green bars would contain blue parts indicating the G>A mutations. **(D)** All WGS samples with coverage below 40 x (n = 6) and 31 additional FFPE CNSL samples (n = 21 PCNSL, n = 10 SCNSL) were analysed by Sanger sequencing. The Sanger sequencing chromatograms showing representative sequences of the *TERT* promoter region. A representative case of an Oligodendroglioma (positive control (CTRL)) showed a C228T mutation and a wild type C250. No *TERT* mutations have been detected in our cohort of CNSL patients (as exemplarily illustrated for patients LS-010 and LS-008).

### References

1. Zhou Y, Zhou B, Pache L, et al: Metascape provides a biologist-oriented resource for the analysis of systems-level datasets. Nat Commun 10(1): 1523; 2019.
