## Supplementary Information for "The genomic and transcriptional landscape of primary central nervous system lymphoma"

### Supplementary material and methods section

#### LymphGen Prediction

The LymphGen algorithm as described by Wright et al.[1] categorises diffuse large B-Cell lymphoma (DLBCL) samples into its different subtypes MCD, N1, A53, BN2 ST2, EZB, $MYC^+$ and $MYC^-$. For this categorization, the algorithm requires three non-optional and three optional files containing information on the mutations and the copy number variations respectively.

- A sample annotation file containing information on processed copy number variation and BCL2 or BCL6 translocation
- A mutation flat file which contains information about nonsense, missense or frameshift mutation in the coding region of the respective gene. Special consideration is given for mutations occurring in the 5'UTR of the respective gene or synonymous mutations in the coding region within 4kb of the transcription start site.
- A list of NCBI Entrez Gene IDs for the for all the genes analysed for mutations
- A file containing the copy number information of various regions, the copy number information is required for prediction of A53 subtype, there are four different types of copy number variation defined for the lymphGen algorithm Gain (single copy number increase), Amp (multiple copy number increase), HETLOSS (Loss of Heterozygousity) and HOMDEL (complete loss of both alleles).
- A file containing the list of NCBI Entrez Gene IDs for the for all the genes analysed for copy number variation
- A file containing information of the impact of a CNV on the chromosome arm.

To create these files, the structural variation workflow Sophia[2] was used for information on BCL2 and BCL6 translocation. The output of SNVCallingWorkflow[3] was used filling the mutations flat file with non-sense or missense mutations, whereas the output of IndelCallingWorkflow[4] was used for annotating the frameshift mutations. ACEseqWorkflow[5] output was processed for extraction of CNVs. The outputs from all workflows were filtered for somatic regions with all different variations occurring in exons and 5'UTR region of the gene. The six files were created for the PCNSL cohort using Python and Perl scripts based on the description provided on the LymphGen website[6]. The individual sample inputs are further merged together to form the input dataset for the LymphGen algorithm and uploaded to the website[7] for classification the samples.

#### Sankey Plot

We further mapped the LymphGen algorithm results to those of the Hans Classifier and RNA-Seq based classification using the adapted figure found in the graphical abstract of Wright et al.[1]

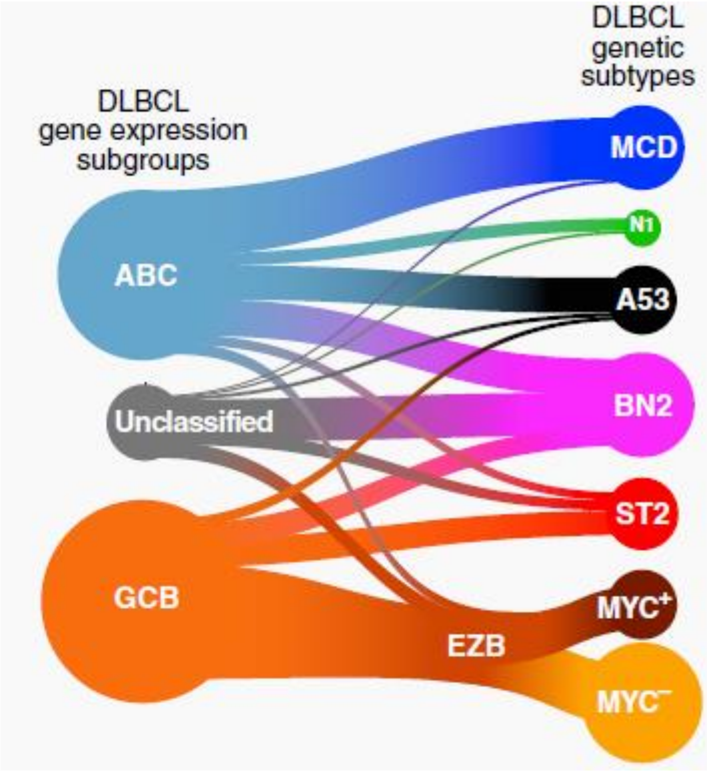


### References


1. Wright GW, Huang DW, Phelan JD, et al: A Probabilistic Classification Tool for Genetic Subtypes of Diffuse Large B Cell Lymphoma with Therapeutic Implications. Cancer Cell 37(4): 551-568 e514; 2020.
3. https://github.com/DKFZ-ODCF/SophiaWorkflow
4. https://github.com/DKFZ-ODCF/SNVCallingWorkflow
5. https://github.com/DKFZ-ODCF/ACEseqWorkflow
6. https://github.com/DKFZ-ODCF/IndelCallingWorkflow
7. https://llmpp.nih.gov/lymphgen/LymphGenInstructions.pdf?v=1600863825
8. https://llmpp.nih.gov/lymphgen/lymphgendataportal.php

### Full list of the member of the MMML consortium

*Coordination (C1):* Reiner Siebert[1,2], Susanne Wagner[2], Andrea Haake[2], Julia Richter[2,3], Gesine Richter[2]
*Data Center (C2):* Roland Eils[4,5], Chris Lawerenz[4], Jürgen Eils[4], Jules Kerssemakers[4], Christina Jaeger-Schmidt[4], Ingrid Scholz[4]
*Clinical Centers (WP1):* Anke K. Bergmann[2, 6], Christoph Borst[7], Friederike Braulke[8], Birgit Burkhardt[9,10], Alexander Claviez[6], Martin Dreyling[11], Sonja Eberth[11], Hermann Einsele[12], Norbert Frickhofen[13], Siegfried Haas[7], Martin-Leo Hansmann[14], Dennis Karsch[15], Nicole Klepl[8], Michael Kneba[15], Jasmin Lisfeld[9], Luisa Mantovani-Löffler[16], Marius Rohde[9], German Ott[17], Christina Stadler[8], Peter Staib[18], Stephan Stilgenbauer[19], Lorenz Trümper[8] , Thorsten Zenz[20]
*Normal Cells (WPN):* Martin-Leo Hansmann[14], Dieter Kube[8], Ralf Küppers[21], Marc Weniger[21]
*Pathology and Analyte Preparation (WP2-3):* Siegfried Haas[7], Michael Hummel[22], Wolfram Klapper[3], Ulrike Kostezka[23], Dido Lenze[22], Peter Möller[24], Andreas Rosenwald[25], German Ott[17], Monika Szczepanowski[3]
*Sequencing and genomics (WP4-7):* Ole Ammerpohl[1,2], Sietse M. Aukema[2,3], Vera Binder[26], Arndt Borkhardt[26], Andrea Haake[2], Jessica I. Hoell[26]; Ellen Leich[25], Peter Lichter[27], Cristina López[1,2], Inga Nagel[2], Jordan Pischimariov[25], Bernhard Radlwimmer[27], Julia Richter[2,3], Philip Rosenstiel[28], Andreas Rosenwald[25], Markus Schilhabel[28], Stefan Schreiber[29], Inga Vater[2], Rabea Wagener[1,2], Reiner Siebert[1,2]
*Bioinformatics (WP8-9):* Stephan H. Bernhart[30-32,] Hans Binder[30,31], Benedikt Brors[33], Gero Doose[30-32], Roland Eils[4,5], Steve Hoffmann[30-32], Lydia Hopp[30], Daniel Hübschmann[4,5,34], Kortine Kleinheinz[4,5], Helene Kretzmer[30-32], Markus Kreuz[35], Jan Korbel[36], David Langenberger[30-32], Markus Loeffler[35], Maciej Rosolowski[35], Matthias Schlesner[4,37], Peter F. Stadler[30-32,38-40], Stephanie Sungalee[36]

[1]Institute of Human Genetics, University of Ulm and University Hospital of Ulm, Ulm, Germany
[2]Institute of Human Genetics, Christian-Albrechts-University, Kiel, Germany;
[3]Hematopathology Section, Institute of Pathology, Christian-Albrechts-University, Kiel, Germany;
[4]Division of Theoretical Bioinformatics (B080), German Cancer Research Center (DKFZ), Heidelberg, Germany;
[5]Department for Bioinformatics and Functional Genomics, Institute of Pharmacy and Molecular Biotechnology and Bioquant, University of Heidelberg, Heidelberg, Germany;
[6]Department of Pediatrics, University Hospital Schleswig-Holstein, Campus Kiel, Kiel, Germany;
[7]Department of Internal Medicine/Hematology, Friedrich-Ebert-Hospital, Neumünster;
[8]Department of Hematology and Oncology, Georg-August-University of Göttingen, Göttingen, Germany;
[9]University Hospital Muenster - Pediatric Hematology and Oncology, Muenster Germany;
[10]University Hospital Giessen, Pediatric Hematology and Oncology, Giessen, Germany;
[11]Department of Medicine III - Campus Grosshadern, University Hospital Munich, Munich, Germany;
[12]University Hospital Würzburg, Department of Medicine and Poliklinik II, University of Würzburg, Würzburg;
[13]Department of Medicine III, Hematology and Oncology, Dr. Horst-Schmidt-Kliniken of Wiesbaden, Wiesbaden;
[14]Senckenberg Institute of Pathology, University of Frankfurt Medical School, Frankfurt am Main, Germany
[15]Department of Internal Medicine II: Hematology and Oncology, University Medical Centre, Campus Kiel, Kiel;
[16]Hospital of Internal Medicine II, Hematology and Oncology, St-Georg Hospital Leipzig, Leipzig, Germany;
[17]Department of Pathology, Robert-Bosch-Hospital, Stuttgart, Germany;
[18]Clinic for Hematology and Oncology, St.-Antonius-Hospital, Eschweiler;
[19]Department for Internal Medicine III, University of Ulm and University Hospital of Ulm, Ulm, Germany
[20]National Centre for Tumor Disease, Heidelberg, Germany;
[21]Institute of Cell Biology (Cancer Research), University of Duisburg-Essen, Duisburg-Essen, Medical School, Essen, Germany;
[22]Institute of Pathology, Charité – University Medicine Berlin, Berlin, Germany;
[23]Comprehensive Cancer Center Ulm (CCCU), University Hospital Ulm, Ulm, Germany;
[24]Institute of Pathology, University of Ulm and University Hospital of Ulm, Ulm;
[25]Institute of Pathology, Comprehensive Cancer Center Mainfranken, University of Wurzburg, Germany;
[26]Department of Pediatric Oncology, Hematology and Clinical Immunology, Heinrich-Heine-University, Düsseldorf, Germany;
[27]German Cancer Research Center (DKFZ), Division of Molecular Genetics, Heidelberg, 69120, Germany;

[28]Institute of Clinical Molecular Biology, Christian-Albrechts-University, Kiel, Germany;
[29]Department of General Internal Medicine, University Kiel, Kiel, Germany;
[30]Interdisciplinary Center for Bioinformatics, University of Leipzig, Leipzig, Germany;
[31]Bioinformatics Group, Department of Computer, University of Leipzig, Leipzig, Germany;
[32]Transcriptome Bioinformatics, LIFE Research Center for Civilization Diseases, University of Leipzig, Leipzig, Germany;
[33]Division of Applied Bioinformatics (G200), German Cancer Research Center (DKFZ), Heidelberg, Germany
[34]Department of Pediatric Immunology, Hematology and Oncology, University Hospital, Heidelberg, Germany
[35]Institute for Medical Informatics Statistics and Epidemiology, University of Leipzig, Leipzig, Germany;
[36]EMBL Heidelberg, Genome Biology, Heidelberg, Germany;
[37]Bioinformatics and Omics Data Analytics (B240), German Cancer Research Center (DKFZ), Heidelberg, Germany;
[38]RNomics Group, Fraunhofer Institute for Cell Therapy and Immunology IZI, Leipzig, Germany
[39]Santa Fe Institute, Santa Fe, New Mexico, United States of America
[40]Max-Planck-Institute for Mathematics in Sciences, Leipzig, Germany.
